# Frailty and loneliness among community-dwelling older adults: Examining reciprocal associations within a measurement burst design

**DOI:** 10.1101/2024.10.07.24314990

**Authors:** Anna Schultz, Hannes Mayerl, Wolfgang Freidl, Erwin Stolz

## Abstract

**Background:** Previous research indicates that frailty and loneliness are interrelated. The aim of this study is to analyze their possible reciprocal relationship while disentangling between- and within-person effects. The separation of these sources of variance is vital for a better understanding of potential causal mechanisms.

**Methods:** Within the FRequent health Assessment In Later life (FRAIL70+) project, participants aged 70 and over completed two measurement bursts spread one year apart with seven biweekly assessments each. The final sample consisted of 426 individuals at baseline (*M*_age_=77.2; *SD*=5.4; 64.6% female). A latent curve model with structured residuals was used to examine the potential reciprocal relationship between frailty (37-item deficit accumulation approach) and loneliness (3-item UCLA scale).

**Results:** No relevant cross-lagged effects over repeated 2-week periods were found between frailty and loneliness at the within-person level, but increases in frailty co-occurred with increases in loneliness. At the between-person level, higher levels of frailty correlated with higher levels of loneliness in each burst.

**Conclusion:** The findings do not support the assumption that frailty and loneliness share a causal reciprocal relationship over weeks and months. Nonetheless, higher levels of frailty were weakly associated with higher levels of loneliness at the within- and considerably associated at the between-person level, which may indicate a common source of both domains.

## Background

Throughout their lives, people may experience different levels of loneliness, which is characterized by an imbalance between their current and ideal level of emotional closeness in social relationships (1,2). Reviews highlight loneliness’s detrimental effects on mental and physical health, including depressive symptoms, sleep quality, decreased immunity, cognitive decline, and mortality (3–5), emphasizing its public health significance (6). Older adults seem to be especially susceptible to loneliness (7), with levels often increasing in later life (1), due to the loss of spouses or health problems (8).

Frailty, an indicator of overall health in older adults (9), increases with age (10), and predicts disability, falls, hospitalization, and mortality (11–13) among older adults. Frailty is common among older adults (14), observed in 12-26% (15), and has increased in more recent birth cohorts (16), highlighting it as a grand challenge (10).

Longitudinal studies showed that high levels of loneliness, on the one hand, increase the risk of developing frailty among older adults (17,18) and on the other, suppress the reversion from (pre-)frailty to robustness (19,20). By contrast, research has also demonstrated that frailty and related health outcomes exert an influence on loneliness. In a sample of *N*=552 Spanish older adults, frailty domains (physical, psychological, and social) predicted loneliness longitudinally (21). Likewise, Hoogendijk et al. (22) found a moderate to large effect (Cohen’s *d* = 0.54) of frailty on loneliness in a sample of *N*=856 older adults in the Netherlands.

While previous research has suggested loneliness as a risk factor for frailty, and frailty as a risk factor for loneliness; a single longitudinal study using cross-lagged panel models also suggested a bidirectional relationship between frailty and loneliness. Using three waves of the China Health and Retirement Longitudinal Study, with a time span of 2 years between waves, Shah and colleagues (23) found among *N =* 2,412 Chinese older adults that frailty and loneliness influence each other, with frailty having a stronger effect on subsequent loneliness than the reverse.

Previous studies have focused on long-term dynamics between loneliness and frailty over multiple years owing to sparse annual or biannual assessment schedules in most health and ageing studies. However, changes likely happen on a much shorter time scale, that is, over weeks and months. For instance, if social contacts of an older adult are severely reduced due to acute or chronic health problems, e.g., an injury that limits mobility or a disability that limits communication, loneliness may follow within weeks or months rather than years. Conversely, a lack of social contacts that usually encourage physical activity, e.g., going for a walk or grocery shopping, could lead to a decline in stamina or muscle strength over the same short-term period.

In the present study, we use data gathered within a measurement burst design to examine the bidirectional relationship between frailty and loneliness over a couple of weeks and months. Understanding these dynamics is crucial, as lonely older adults living with frailty have a higher mortality risk compared to those who are only frail or lonely (24). We attempt to shed light onto the frailty-loneliness interplay by distinguishing associations existing at the within-person level (i.e., whether a person experiencing lower health than usual also reports higher loneliness at the same or adjacent point in time) from those at the between-person level (i.e., whether individuals with greater health declines than others are also more likely to experience greater loneliness). This distinction is important, as within- and between-person associations can differ considerably in magnitude or direction (25), highlighting its significance for understanding cause-effect relationships in longitudinal observational data (26).

## Methods

### Design and Data

In the FRequent health Assessment In Later life (FRAIL70+) project, nationwide health data of Austrian community-dwelling older adults aged 70 and above were collected between August 2021 and April 2023 (Supplementary Methods 1; Supplementary Figure 1) using a measurement burst design (27). This design involves conducting several assessments (e.g., performance tests, standardized questionnaires) within a relatively short time span (biweekly), which are then repeated at longer intervals (annually). In the first burst, 426 participants (response rate: 44%) completed up to seven biweekly interviews (retention rate=95.3-98.5%). One year later, 378 participants returned for a second burst (retention rate between bursts=88.7%), consisting again of seven biweekly assessments (retention rate= 76.7-94.2%; last interview=53.5%). The study was approved by the Ethics Committee of the Medical University of Graz (EK-number: 33-243 ex 20/21). All participants provided informed consent.

## Measures

### Frailty

Frailty was operationalized using the Frailty Index (FI; 28), one of the primary methods for assessing frailty (10), through various self-reported health items and cognitive performance tests (29). A FI was computed from 37 items (see Supplementary Table 1 for more details). No item had > 1.7% missing values. Each item was first mapped to the interval of 0-1 (i.e., dichotomous items were scored 0 or 1, categorical items had, for instance, scores of 0, 0.25, 0.5, 0.75, or 1.0) and then, provided that at least 80% of data were valid, summed up and divided by the number of valid health deficits (e.g., 11/37=0.23), resulting in a score ranging (theoretically) between 0 and 1, whereby higher values indicate higher frailty levels. Internal consistency in the current study was ω=0.89 and is in line with previous studies (ω=.89-.93 (29); ω=.81 (30)).

### Loneliness

Loneliness (LS) was measured using the three-item University of California, Los Angeles loneliness scale (UCLA; 32; see Supplementary Table 2 for more details). LS scores range from 3 to 9 with higher scores indicating higher levels of LS. Previous research reports satisfactory reliability (α=0.72) as well as concurrent and discriminant validity (32). Internal consistency in the current study was ω=0.76.

### Additional Variables

Baseline variables for multiple group analysis, assessed during the first interview in the first burst, included sex (male vs. female), age groups (70-74, 75-79, vs. >80 years), living alone (no vs. yes), and social participation (no vs. yes). The FRAIL70+ survey asked participants whether they participated in the following social activities during the past year: (1) volunteer or charitable work, (2) participation in religious institutions (e.g., church, synagogue, or mosque), (3) participation in political organizations or citizens’ initiatives, and (4) participation in educational or training courses. Responses were summed and categorized as 0=“no” and 1=“yes, participation in one or more activities”.

## Statistical Analysis

Following preliminary descriptive analyses, we employed latent curve modeling with structured residuals (LCM-SR; 33) using maximum likelihood estimation with robust (Huber-White) standard errors (MLR) and full information maximum likelihood (FIML) estimation to address missing data. In the Supplementary Methods 2, we outline the advantage of the latent curve over the mixed effects modeling framework. Unlike cross-lagged panel models, where between- and within-effects are intermingled, LCM-SR separates between- and within-person effects. This is accomplished by combining the cross-lagged panel model with the latent growth curve model, and by modeling autoregressive, within-time and cross-lagged effects among the time-specific residuals (33). Within the LCM-SR, latent intercept and slope factors for FI and LS are specified to reflect between-person variability. Within-person effects then account for the remaining variability in each observation after modelling the between-person intercept and slope. In other words, within-person cross-lagged effects refer to how individual deviations (represented by the time-specific residuals) from a person’s trajectory (represented by the random intercept and slope) in one domain (e.g., the FI) are related to concurrent and subsequent deviations in another domain (e.g., LS). While between-person effects acknowledge that individuals may differ in both their levels and growth over time due to factors, like gender or educational level, these effects can be confounded with various background characteristics, including genetics, early-life factors, personality, and demographic factors. Within-person effects, on the other hand, control for such stable (un)observable time-invariant confounders (34). This is a significant advantage of the LCM-SR, addressing a key challenge in non-experimental study designs where controlling for all relevant covariates is not possible, hindering causal effect estimation. Taking a within-person analytical approach can be seen as a step towards uncovering causal effects, as it enables the identification of individual-level dynamics that might otherwise be masked by group-level trends (35).

Statistical analysis comprised three stages. First, we established the optimally fitting model for each outcome variable separately, comparing four models with increasing complexity: (1) a model without any random effects (i.e., only autoregressive effects; e.g., past FI predicts current FI), and subsequently added (2) random intercepts, (3) fixed slopes, and (4) random slopes. Thereby, individuals were allowed to vary around their person-specific levels (model 2) and around their person-specific trajectories (model 4) rather than group-level averages. In all models, we specified the residuals of the observed variables as latent, and added autoregressive paths between adjacent residuals (α, β). To account for the heterogeneous measurement occasions owing to the measurement burst design, we specified one intercept and one slope for each burst (for a similar approach see 35). Model comparison was based on likelihood ratio tests (37).

Second, we combined the best-fitting univariate models to build the bivariate model. This involved adding cross-lagged regression paths from latent residuals in one construct at a specific time to the subsequent latent residuals in the other construct (γ, δ), and allowing latent residuals across constructs to covary within-time (λ). Substantial and statistically significant cross-lagged effects would provide evidence for a causal relationship between the frailty and loneliness, while covarying residuals would instead suggest a common cause of both constructs. A graphical representation of the final model can be found in Figure 1. Third, we conducted multiple group analysis for sex, age, living alone, and social participation.

**Figure 1:**
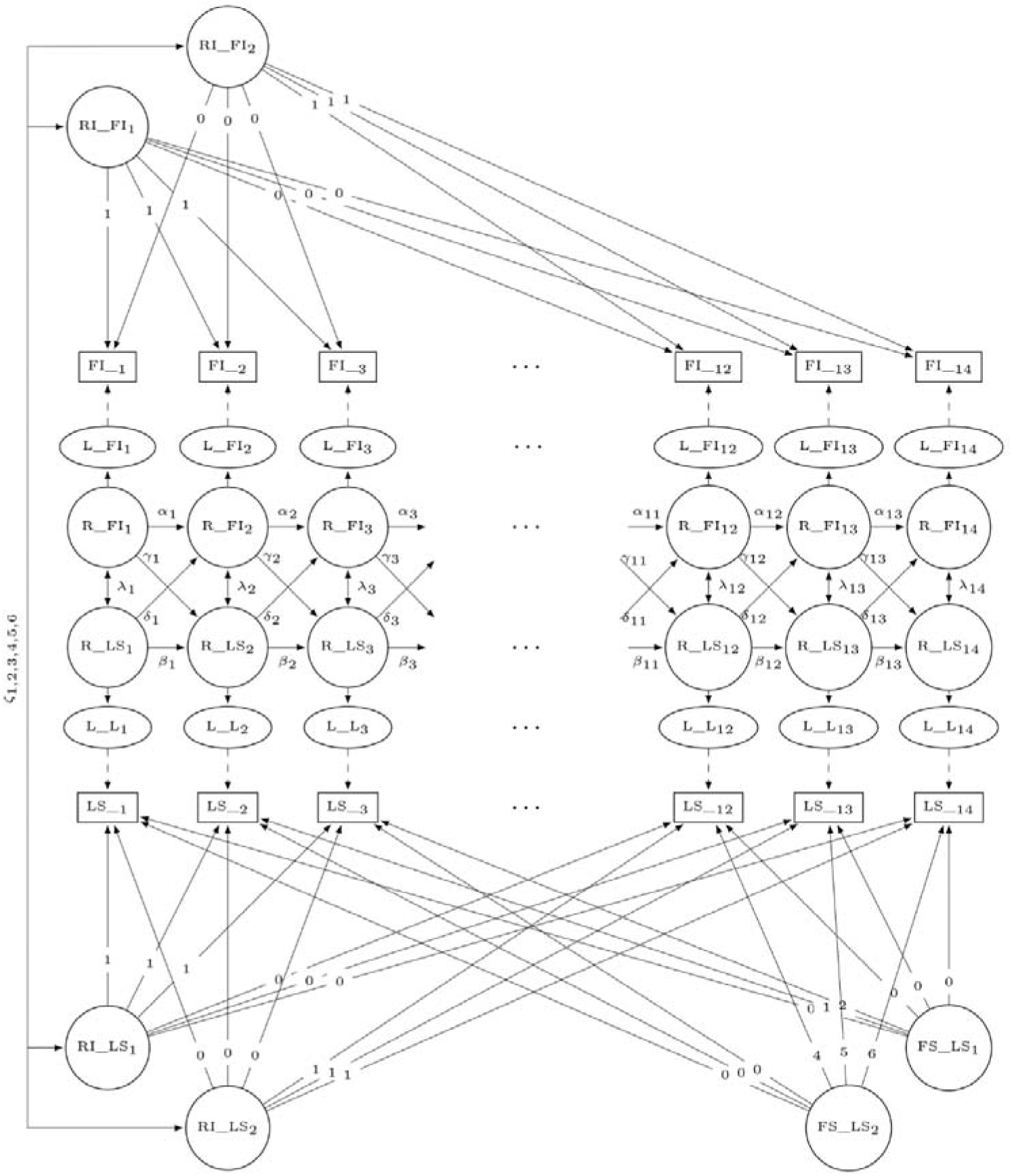
Bivariate latent curve model with structured residuals (LCM-SR) for the frailty index (FI) and loneliness (LS) across waves 1-14. Manifest variables (= computed scores) are represented as rectangles, latent variables as ellipses, and residuals as circles. Single and double headed arrows denote regression paths and covariances, respectively. Paths labeled with numbers index the coding of the intercept and linear slope. Greek symbols indicate that the respective path is estimated freely. RI = random intercept. FS = fixed slope. L = latent variable. R = residual.

Model fit was evaluated using robust variants of several fit indices: Tucker-Lewis index (TLI), comparative fit index (CFI), standardized root mean square residual (SRMR), and root mean square error of approximation (RMSEA) with 90% confidence interval. Adequate fit was defined as CFI and TLI ≥ 0.95, and RMSEA and SRMR ≤ 0.06 (38). Statistical analyses were carried out in R 4.2.3 (39) using the lavaan (40, version 0.6-15) and semTools (41, version 0.5-6) packages.

## Results

### Sample Characteristics

The average age of participants at baseline was 77.0 years (*SD*=5.4), 64.6% were female, and 66.0% were living alone. The majority (54.2%) reported a medium level of education, 19.2% reported a low, and 26.5% a high level of education (i.e., A-levels or a university degree). At baseline, 41.5% of participants indicated to take part in at least one social activity, whereby attending clubs emerged as the most frequently selected activity (26.8%). Mean (*SD*) and median (*IQR*) of baseline FI and LS were 0.2 (*SD*=0.1) and 0.1 (*IQR*=0.2), and 3.5 (*SD*=1.1) and 3.0 (*IQR*=1.0), respectively. The majority of older adults was not frail (67.1% based on the cut-off of the FI ≤0.2; 41) and not lonely (94.3% based on the cut-off of LS score ≤5; 42). Participants completed a median of 13 (IQR = 3.0, range=1-14) interviews. Additional descriptive statistics of the FI and LS by measurement occasion are shown in Table 1. Pairwise correlations showed a positive association between the FI and LS (see Supplementary Figure 2), which on average amounted to *r*=0.38 (range=0.17-0.50). Within construct mean correlation coefficients across waves were *r*=0.83 (range=0.71-0.94) and *r*=0.64 (range=0.44-0.87) for the FI and LS, respectively.

**Table 1:** Descriptive statistics of frailty and loneliness by wave.

| Wave | *n* | FI |  | LS |  |
| --- | --- | --- | --- | --- | --- |
|  |  | Mean (*SD*) | Median (*IQR*) | Mean (*SD*) | Median (*IQR*) |
| 1 | 426 | 0.19 (0.14) | 0.15 (0.16) | 3.51 (1.08) | 3 (1) |
| 2 | 419 | 0.18 (0.14) | 0.14 (0.15) | 3.40 (0.96) | 3 (0) |
| 3 | 419 | 0.17 (0.14) | 0.14 (0.14) | 3.36 (0.94) | 3 (0) |
| 4 | 410 | 0.18 (0.14) | 0.14 (0.16) | 3.38 (0.92) | 3 (0) |
| 5 | 406 | 0.17 (0.13) | 0.14 (0.14) | 3.42 (0.95) | 3 (0) |
| 6 | 407 | 0.18 (0.13) | 0.14 (0.15) | 3.49 (1.01) | 3 (1) |
| 7 | 406 | 0.18 (0.13) | 0.15 (0.15) | 3.55 (1.06) | 3 (1) |
| 8 | 378 | 0.20 (0.15) | 0.15 (0.16) | 3.49 (1.01) | 3 (1) |
| 9 | 350 | 0.19 (0.14) | 0.16 (0.16) | 3.44 (0.96) | 3 (1) |
| 10 | 356 | 0.19 (0.14) | 0.15 (0.16) | 3.43 (0.98) | 3 (0) |
| 11 | 334 | 0.19 (0.13) | 0.15 (0.16) | 3.55 (1.12) | 3 (1) |
| 12 | 320 | 0.19 (0.14) | 0.15 (0.18) | 3.37 (0.86) | 3 (0) |
| 13 | 290 | 0.20 (0.14) | 0.16 (0.18) | 3.46 (1.02) | 3 (0) |
| 14 | 202 | 0.19 (0.14) | 0.16 (0.16) | 3.47 (1.15) | 3 (0) |

*Note.* FI = frailty index, LS = loneliness, *SD* = standard deviation; *IQR* = interquartile range.

Those who dropped out in any wave (N = 264) were not frailer (M_1_ = 0.18, SD_1_ = 0.12 vs. M_2_ = 0.20, SD_2_ = 0.15; *t* = −1.602, *p*=.110) or lonelier (M_1_ = 3.40, SD_1_ = 0.85 vs. M_2_ = 3.57, SD_2_ = 1.19; Mann-Whitney-*U* = 20412, *p*=.476) at baseline compared to the remaining sample. No significant differences were found (p<.05) with regard to socio-demographic characteristics, i.e., sex, age, education, living situation.

### Univariate models

Parameters and model fit statistics of univariate models for the FI and LS are displayed in Supplementary Tables 3-6. We followed the model building strategy outlined in the method section, starting with a model without any random effects and gradually increasing complexity by adding random intercept and slope factors. This approach reflects previous findings that baseline levels and rates of change vary between individuals (e.g., 1,10,44).

For both constructs, model 1 (i.e., a model without any random effects) fit poorly, but adding random intercepts significantly improved model fit (model 1 vs. model 2). For the FI, model fit did not improve further when fixed slopes were added (model 2 vs. model 3), therefore, we did not increase complexity through the inclusion of a random slope (Supplementary Table 4). In contrast, for LS, adding fixed slopes did significantly improve model fit. However, including a random slope did not result in better model fit (model 3 vs. model 4; Supplementary Table 6). Based on the results of the univariate models, we combined the FI model 2 (i.e., random intercepts) with LS model 3 (i.e., random intercepts and fixed slopes) to create the bivariate model.

For the final FI model, we found mostly positive autoregressive effects of small size (see Supplementary Table 3). The model implied intercepts amounted to 0.18 (95% CI=0.17, 0.19) and 0.20 (95% CI=0.19, 0.22) for the first and second burst, respectively. Correlation of intercepts was very high (ζ= 0.94, 95% CI=0.92, 0.97), indicating that older adults with higher frailty levels in the first burst showed also higher frailty levels in the second burst.

Autoregressive parameters for the final LS model, i.e., the within-person stability of LS, tended to show positive, yet weak effects (see Supplementary Table 5). The model implied intercepts amounted to 3.39 (95% CI=3.30, 3.48) and 3.51 (95% CI=3.41, 3.61) for the first and second burst, respectively. Average LS changed minimally within bursts and only little between bursts. Correlation of intercepts was again very high (ζ=0.92; 95% CI=0.86, 0.98), indicating that participants who were lonelier in the first burst were also lonelier in the second.

### Bivariate model

The final LCM-SR yielded good model fit: χ^2^(324)=700.39, *p*< .001; CFI=0.986, TLI=0.983; SRMR=0.049; RMSEA=0.042, 90% CI=0.029, 0.053. Parameter estimates are shown in Table 2.

**Table 2:** Parameter estimates of the bivariate model.

| Parameter | Est. [95% CI] |
| --- | --- |
| Random effects: Means |  |
| Intercept FI1<sup>*</sup> | 0.18 [0.17, 0.19] |
| Intercept FI2<sup>*</sup> | 0.20 [0.19, 0.22] |
| Intercept LS1<sup>*</sup> | 3.39 [3.30, 3.48] |
| Intercept LS2<sup>*</sup> | 3.51 [3.42, 3.61] |
| Fixed effects: Means |  |
| Slope LS1<sup>*</sup> | 0.02 [0.00, 0.03] |
| Slope LS2<sup>*</sup> | -0.01 [-0.02, 0.00] |
| Random Effects: Correlation |  |
| $\zeta_1$: Intercept FI1 ↔ Intercept FI2 | 0.94 [0.92, 0.97] |
| $\zeta_2$: Intercept FI1 ↔ Intercept LS1 | 0.57 [0.46, 0.68] |
| $\zeta_3$: Intercept FI1 ↔ Intercept LS2 | 0.58 [0.46, 0.69] |
| $\zeta_4$: Intercept FI2 ↔ Intercept LS1 | 0.52 [0.41, 0.63] |
| $\zeta_5$: Intercept FI2 ↔ Intercept LS2 | 0.53 [0.41, 0.65] |
| $\zeta_6$: Intercept LS1 ↔ Intercept LS2 | 0.92 [0.87, 0.98] |
| Autoregressive (FI → FI) | |
| $\alpha_1$ | 0.26 [0.11, 0.41] |
| $\alpha_2$ | 0.29 [0.15, 0.43] |
| $\alpha_3$ | 0.28 [0.04, 0.53] |
| $\alpha_4$ | 0.08 [-0.10, 0.26] |
| $\alpha_5$ | 0.28 [0.10, 0.45] |
| $\alpha_6$ | 0.21 [0.04, 0.38] |
| $\alpha_7$ | 0.12 [-0.13, 0.37] |
| $\alpha_8$ | 0.23 [0.07, 0.38] |
| $\alpha_9$ | 0.17 [-0.02, 0.36] |
| $\alpha_{10}$ | 0.11 [-0.23, 0.45] |
| $\alpha_{11}$ | 0.29 [0.07, 0.52] |
| $\alpha_{12}$ | 0.22 [0.02, 0.41] |
| $\alpha_{13}$ | 0.34 [0.08, 0.59] |
| Autoregressive (LS → LS) | |
| $\beta_1$ | 0.24 [0.08, 0.39] |
| $\beta_2$ | 0.16 [-0.07, 0.39] |
| $\beta_3$ | 0.14 [-0.13, 0.40] |
| $\beta_4$ | 0.03 [-0.21, 0.28] |
| $\beta_5$ | 0.14 [-0.09, 0.37] |
| $\beta_6$ | 0.26 [0.09, 0.44] |
| $\beta_7$ | -0.02 [-0.28, 0.24] |
| $\beta_8$ | 0.07 [-0.16, 0.29] |
| $\beta_9$ | 0.06 [-0.23, 0.35] |
| $\beta_{10}$ | 0.03 [-0.24, 0.29] |
| $\beta_{11}$ | -0.14 [-0.40, 0.12] |
| $\beta_{12}$ | 0.04 [-0.21, 0.29] |
| $\beta_{13}$ | 0.34 [0.07, 0.61] |
| Cross-lagged (LS → FI) | |
| $\delta_1$ | 0.00 [-0.16, 0.16] |
| $\delta_3$ | 0.09 [-0.04, 0.21] |
| $\delta_3$ | 0.05 [-0.10, 0.20] |
| $\delta_4$ | -0.03 [-0.17, 0.12] |
| $\delta_5$ | -0.05 [-0.19, 0.10] |
| $\delta_6$ | -0.03 [-0.15, 0.09] |
| $\delta_7$ | 0.08 [-0.05, 0.20] |
| $\delta_8$ | 0.01 [-0.17, 0.19] |
| $\delta_9$ | 0.03 [-0.15, 0.22] |
| $\delta_{10}$ | -0.02 [-0.20, 0.16] |
| $\delta_{11}$ | 0.09 [-0.06, 0.25] |
| $\delta_{12}$ | 0.04 [-0.12, 0.20] |
| $\delta_{13}$ | 0.08 [-0.08, 0.25] |
| Cross-lagged (FI → LS) | |
| $\gamma_1$ | 0.10 [0.00, 0.20] |
| $\gamma_3$ | 0.16 [-0.01, 0.32] |
| $\gamma_3$ | 0.17 [0.00, 0.34] |
| $\gamma_4$ | -0.07 [-0.26, 0.12] |
| $\gamma_5$ | -0.04 [-0.20, 0.13] |
| $\gamma_6$ | -0.08 [-0.20, 0.04] |
| $\gamma_7$ | 0.05 [-0.07, 0.17] |
| $\gamma_8$ | 0.11 [-0.07, 0.29] |
| $\gamma_9$ | -0.00 [-0.20, 0.19] |
| $\gamma_{10}$ | 0.07 [-0.14, 0.28] |
| $\gamma_{11}$ | 0.23 [0.01, 0.45] |
| $\gamma_{12}$ | 0.05 [-0.17, 0.27] |
| $\gamma_{13}$ | 0.14 [-0.04, 0.33] |
| Within-time (FI ↔ LS) | |
| $\lambda_1$ | 0.23 [0.12, 0.34] |
| $\lambda_2$ | 0.17 [0.05, 0.28] |
| $\lambda_3$ | 0.18 [0.08, 0.29] |
| $\lambda_4$ | 0.14 [-0.03, 0.31] |
| $\lambda_5$ | 0.16 [-0.04, 0.37] |
| $\lambda_6$ | 0.18 [0.03, 0.34] |
| $\lambda_7$ | 0.19 [0.07, 0.32] |
| $\lambda_8$ | 0.07 [-0.08, 0.22] |
| $\lambda_9$ | 0.05 [-0.13, 0.23] |
| $\lambda_{10}$ | 0.08 [-0.08, 0.24] |
| $\lambda_{11}$ | 0.10 [-0.08, 0.28] |
| $\lambda_{12}$ | 0.10 [-0.18, 0.37] |
| $\lambda_{13}$ | 0.11 [-0.02, 0.25] |
| $\lambda_{14}$ | 0.09 [-0.06, 0.24] |

*Note.* We report standardized parameter estimates and 95% CI for all variables, except for the means of random and fixed effects. The superscript \* indicates that unstandardized estimates are reported instead.

As regards the cross-lagged structure, we found weak and mostly positive effects between the FI and LS. Although cross-lagged effects from the FI to LS were greater in magnitude compared to those from LS to the FI, both these within-person effects were negligible in size and the majority of confidence intervals included zero.

Concerning the within-time associations, we found positive, albeit small effects between the FI and LS. That is, individuals who were frailer than usual tended to be also lonelier than usual.

As regards the between-person effects, model implied intercepts amounted to 0.18 (95% CI=0.17, 0.19) and 0.20 (95% CI=0.19, 0.22) for the FI, and to 3.39 (95% CI=3.30, 3.48) and 3.51 (95% CI=3.42, 3.61) for LS. Average LS changed minimally within bursts and only little between bursts. Moreover, random intercepts of the FI and LS share a high positive relationship, indicating that frailer older adults were on average also lonelier.

### Multiple group analysis

Multiple group analysis in terms of sex (men/women), age (70-74 vs. 75-79 vs. >80), living alone (no/yes), and social participation (no/yes) was performed on the final bivariate model. In terms of sex, women showed slightly stronger effects than men in the within-person parameters (see Supplementary Table 7). As regards the between-person effects, we found differences in random intercept factors for both FI and LS: women were both frailer and lonelier at baseline.

Concerning age, living situation, and social participation, within-person effects did not differ notably (see Supplementary Tables 8-10). As anticipated, LS and frailty increased with age: Individuals older than 80 years exhibited higher values in the FI and LS compared to younger participants. Between-person effects also differed with regards to living situation: Individuals living alone were frailer and lonelier compared to individuals not living alone. In terms of social participation, between person effects also showed differences: Individuals who participated in at least one activity in the past year were less frail and less lonely.

## Discussion

The present study aimed at describing the relationship between frailty and loneliness in older adults. It extends previous research by (1) studying short-term dynamics and (2) disentangling between-from within-person effects (i.e., differences manifesting between individuals from processes happening within individuals). The importance of the latter is already recognized in the literature (45) as it allows for studying how changes in one domain predict changes in another at the at the level of the individual (35).

Using intensive longitudinal data from community-dwelling Austrian older adults, we found that an increase in frailty within a person at a specific point in time is *not* associated with an increase in loneliness within that person at a later point in time, and vice versa. Although point estimates for the cross-lagged parameters of frailty on later loneliness are higher compared to those of loneliness on frailty, their practical relevance is negligible. Thus, these findings do not support a direct link between frailty and later loneliness or between loneliness and later frailty. The slightly greater influence frailty exerts on subsequent loneliness contrasts with findings from a recent study (46), which suggests that changes in an older adult’s health status contribute only minimally to feelings of loneliness.

Second, we found a positive within-time relationship between frailty and loneliness. In other words, we found that within-person increases in frailty coincide with worsening levels of loneliness at the same point in time. While the existence of short-term fluctuations in both frailty and loneliness align with previous studies (31,47,48), our findings show that frailty and loneliness go up and down together. These fluctuations may be explained by discrete health-related events, such as infections or injuries: For instance, when an older person falls or catches a cold and becomes (temporarily) bedridden or housebound, they are less able to participate in social life; we would expect a decrease in physical health (= increase in frailty) and an increase in loneliness. To examine whether bedrest or falls (serving as proxies for infections and/or injuries) influence the small within-time relationship between frailty and loneliness, we included both proxies separately (as time-varying covariates) in our final model as a supplementary analysis (see Supplementary Table 11 and 12). This additional analysis revealed that the temporal relationship between frailty and loneliness decreased when these time-varying covariates were included, which suggests that acute health-related events promote both frailty and loneliness, for example via increased pain, fatigue or decreased mobility.

Third, regarding the between-person effects, we found that older adults who show higher levels of frailty also demonstrate higher levels of loneliness at burst one and two, respectively. Taken together, the presence of strong intercept-intercept correlations across constructs along with the absence of cross-lagged within-person relations questions previous findings (23,49,50) of a causal bidirectional relationship between frailty and loneliness.

Comparison with previous results proves difficult, however, since the majority of studies conducted in this context either examined the associations separately (e.g., 19), used different health outcomes (e.g., self-rated health; 49,50), different operationalizations of frailty and loneliness (23), investigated long-term dynamics (over the course of years; e.g., 23), or did not disentangle between-person variability from changes happening within the individual (23,49,50). The latter is vital when examining change processes over a period of time as the collected longitudinal data contains information on within- and between-person effects. Traditional cross-lagged panel modeling assumes that individuals vary around the same means as time unfolds and disregards the possibility of trait-like individual differences, hence “it follows that many lagged parameters reported in the literature will not reflect the actual within-person (causal) mechanisms” (25). Thus, although previous studies found cross-lagged effects between either frailty or alternative health outcomes and loneliness, we have to keep in mind that these effects intermingled between-person differences and within-person changes, which in turn renders them vulnerable to confounding. Cross-lagged regressions within the LCM-SR, however, separate between- and within-effects, and assess whether higher-than-usual frailty predicts higher-than-usual loneliness (or vice-versa). Yet, examining both levels (i.e., within- and between-person effects) enriches our understanding of the interplay between mental and physical health at an advanced age: While within-person effects allow the study of personal dynamics, between-person effects allow the study of variation between individuals and the identification of common patterns and disparities.

Possible reasons for not identifying substantial cross-lagged effects between frailty and loneliness include measurement accuracy. Not directly observable qualities like frailty and loneliness need to be scaled in order to evaluate changes. Yet, the evaluation of changes in frailty and loneliness requires that the scale produces reliable estimates of an individual’s underlying true score. While recent research (30,31,51) demonstrates the reliability of the FI and suggests that it is possible to distinguish between frail and robust individuals (between-person effects), one study (31) indicates that the relatively large standard error of measurement (SEM=0.05) and smallest detectable change (SDC=0.13) render it difficult to monitor within-person changes reliably. As regards the UCLA loneliness scale employed in the current study, previous studies demonstrated internal consistency (see 52) comparable to our findings (ω=0.76 vs. α=0.67-0.88). There is, however, little evidence in terms of test-retest reliability. Although, five studies found adequate test-retest reliability of the full UCLA loneliness scale, no estimates are available for the economic three-item version (53) used in many longitudinal studies on health and ageing (e.g., the English Longitudinal Study of Ageing) and also in the current study.

On the other hand, previous studies reported an association between fatigue, depression, cognitive impairment, and loneliness at an advanced age (54,55). Moreover, lonely older adults have been shown to exercise or move less, suffer from malnutrition (56), and experience functional limitations or depressive symptoms (57), characteristics that are closely related to and indeed often included in the frailty index. In the same vein, loneliness has also been cited as a social component frequently included in frailty assessment instruments (58). In addition to that, recent findings (30,59) suggest that depression and frailty, and depression and loneliness demonstrate a shared vulnerability. It seems therefore possible that the association between frailty and loneliness may also be due to common causes, e.g., lack of social support or social buffering (e.g., 60–62). Having found that the random intercepts and the respective correlations, that is, between-person effects, vary by demographic characteristics (i.e., gender, age, living situation, and social involvement), and that frailty and loneliness fluctuate jointly support this assumption.

To the best of the authors’ knowledge, this is the first study examining the bidirectional relationship between frailty and loneliness while also disentangling within- and between-person effects. Further strengths of the current study include the data quality and study design. We analyzed intensive longitudinal data of a nation-wide cohort of community-dwelling older adults gathered within a measurement burst design. Nevertheless, this study also has limitations. First, the outcomes used in this study (FI and LS) were based almost exclusively on self-reports. In terms of loneliness, this may have led to an underestimation of its prevalence since loneliness comprises an undesirable and often stigmatized emotional state. We tried to overcome this limitation by using an indirect measurement of loneliness where items do not include the term loneliness. Our FI, on the other hand, included only two objective measures (cognitive tests). As the result of a previous study (63) showed that frailty levels were lower when based solely on self-reports, future research could employ FIs that include more test-based health measures. Second, selection effects and sample attrition were present. In particular, younger, female and well-educated individuals were overrepresented in the sample (31). Additionally, more than half of the participants were lost during the second burst, although on average, participants provided 13 (out of 14 possible) repeated measurements. We tried to overcome sample attrition this by employing FIML, a procedure considering all available information about an individual (64).

## Conclusion

Disentangling within- and between-person effects is useful when examining the relationship between frailty and LS, and provides the opportunity to explore the potential reciprocal relationship between these two domains in later life. While there was scarce evidence in our study to support such a direct association, it did suggest that frailty and LS may share short- and long-term causes.

## Data Availability

The R-code to reproduce the statistical analyses are available online via the OSF repository (https://osf.io/63c42/). The Scientific Use File (SUF edition) (65) of the FRAIL70+ data is upon registration available at the Austrian Social Science Data Archive AUSSDA (https://doi.org/10.11587/DJNOHX) for researches and students via download.

https://osf.io/63c42/

## List of Abbreviation

FRAIL70+: FRequent health Assessment In Later life project
UCLA: University of California, Los Angeles loneliness scale
FI: Frailty Index
LS: Loneliness
LCM-SR: Latent growth model with structured residuals
MLR: Maximum likelihood estimation with robust (Huber-White) standard errors
FIML: Full information maximum likelihood estimation
TLI: Robust Tucker-Lewis index
CFI: Robust comparative fit index
SRMR: Robust standardized root mean square residual
RMSEA: Robust root mean square error of approximation

## Declarations

### Ethics Approval and consent to participate

The study was approved by the Ethics Committee of the Medical University of Graz (EK-number: 33-243 ex 20/21). Informed consent was obtained from all participants.

### Consent for publication

Not applicable.

### Availability of data and materials

The R-code to reproduce the statistical analyses are available online via the OSF repository (https://osf.io/63c42/). The Scientific Use File (SUF edition) (65) of the FRAIL70+ data is upon registration available at the Austrian Social Science Data Archive – AUSSDA (www.aussda.at) for researches and students via download.

### Competing interests

The authors declare that they have no competing interests.

### Funding

Data collection of the FRAIL70+ study was funded with a grant from the Austrian Science Fund (FWF [grant number: P33673]).

### Authors’ contributions

ES, HM, and AS conceptualized the study and were involved in the design of the methodology. AS performed all statistical analysis and wrote the first draft of the manuscript. ES, HM and WF critically reviewed the manuscript. All authors read and approved the final manuscript.

## Acknowledgements

Not applicable

## Notes

### Competing Interest Statement

The authors have declared no competing interest.

### Funding Statement

This study was funded by the Austrian Science FUnd (FWF [grant number: P33673]).

### Author Declarations

Ethics committee of Medical University of Graz gave ethical approval of this work (EK-number: 33-243 ex 20/21)

## References

1. Luhmann M, Hawkley LC. Age differences in loneliness from late adolescence to oldest old age. Dev Psychol. 2016 Jun;52(6):943–59.

2. Peplau LA, Perlman D. Loneliness: A sourcebook of current theory, research, and therapy. John Wiley & Sons; 1982.

3. Hawkley LC, Capitanio JP. Perceived social isolation, evolutionary fitness and health outcomes: a lifespan approach. Philos Trans R Soc B Biol Sci. 2015 May 26;370(1669):20140114.

4. Holt-Lunstad J, Smith TB, Baker M, Harris T, Stephenson D. Loneliness and Social Isolation as Risk Factors for Mortality: A Meta-Analytic Review. Perspect Psychol Sci. 2015 Mar;10(2):227–37.

5. Wang F, Gao Y, Han Z, Yu Y, Long Z, Jiang X, et al. A systematic review and meta-analysis of 90 cohort studies of social isolation, loneliness and mortality. Nat Hum Behav. 2023 Jun 19;7(8):1307–19.

6. Holt-Lunstad J. Social Connection as a Public Health Issue: The Evidence and a Systemic Framework for Prioritizing the “Social” in Social Determinants of Health. Annu Rev Public Health. 2022 Apr 5;43(1):193–213.

7. Victor CR, Yang K. The Prevalence of Loneliness Among Adults: A Case Study of the United Kingdom. J Psychol. 2012 Jan;146(1–2):85–104.

8. Dahlberg L, McKee KJ, Frank A, Naseer M. A systematic review of longitudinal risk factors for loneliness in older adults. Aging Ment Health. 2022 Feb 1;26(2):225–49.

9. Rockwood K, Mitnitski A. Frailty in Relation to the Accumulation of Deficits. J Gerontol A Biol Sci Med Sci. 2007 Jul 1;62(7):722–7.

10. Hoogendijk EO, Afilalo J, Ensrud KE, Kowal P, Onder G, Fried LP. Frailty: implications for clinical practice and public health. The Lancet. 2019 Oct;394(10206):1365–75.

11. Kojima G, Iliffe S, Walters K. Frailty index as a predictor of mortality: a systematic review and meta-analysis. Age Ageing. 2018 Mar 1;47(2):193–200.

12. Kojima G, Taniguchi Y, Aoyama R, Tanabe M. Associations between loneliness and physical frailty in community-dwelling older adults: A systematic review and meta-analysis. Ageing Res Rev. 2022 Nov;81:101705.

13. Vermeiren S, Vella-Azzopardi R, Beckwée D, Habbig AK, Scafoglieri A, Jansen B, et al. Frailty and the Prediction of Negative Health Outcomes: A Meta-Analysis. J Am Med Dir Assoc. 2016 Dec;17(12):1163.e1–1163.e17.

14. O’Caoimh R, Galluzzo L, Rodríguez-Laso Á, Van der Heyden J, Ranhoff AH, Lamprini-Koula M, et al. Prevalence of frailty at population level in European ADVANTAGE Joint Action Member States: a systematic review and meta-analysis. Ann Ist Super Sanita. 2018;54(3):226–38.

15. O’Caoimh R, Sezgin D, O’Donovan MR, Molloy DW, Clegg A, Rockwood K, et al. Prevalence of frailty in 62 countries across the world: a systematic review and meta-analysis of population-level studies. Age Ageing. 2021 Jan 8;50(1):96–104.

16. Hoogendijk EO, Stolz E, Oude Voshaar RC, Deeg DJH, Huisman M, Jeuring HW. Trends in Frailty and Its Association With Mortality: Results From the Longitudinal Aging Study Amsterdam, 1995–2016. Am J Epidemiol. 2021 Jul 1;190(7):1316–23.

17. Davies K, Maharani A, Chandola T, Todd C, Pendleton N. The longitudinal relationship between loneliness, social isolation, and frailty in older adults in England: a prospective analysis. Lancet Healthy Longev. 2021 Feb;2(2):e70–7.

18. Ge L, Yap CW, Heng BH. Associations of social isolation, social participation, and loneliness with frailty in older adults in Singapore: a panel data analysis. BMC Geriatr. 2022 Dec;22(1):26.

19. Jarach CM, Tettamanti M, Nobili A, D’avanzo B. Social isolation and loneliness as related to progression and reversion of frailty in the Survey of Health Aging Retirement in Europe (SHARE). Age Ageing. 2021 Jan 8;50(1):258–62.

20. Sha S, Xu Y, Chen L. Loneliness as a risk factor for frailty transition among older Chinese people. BMC Geriatr. 2020 Dec;20(1):300.

21. Doñate-Martínez A, Alhambra-Borrás T, Durá-Ferrandis E. Frailty as a Predictor of Adverse Outcomes among Spanish Community-Dwelling Older Adults. Int J Environ Res Public Health. 2022 Oct 5;19(19):12756.

22. Hoogendijk EO, Suanet B, Dent E, Deeg DJH, Aartsen MJ. Adverse effects of frailty on social functioning in older adults: Results from the Longitudinal Aging Study Amsterdam. Maturitas. 2016 Jan;83:45–50.

23. Sha S, Pan Y, Xu Y, Chen L. Associations between loneliness and frailty among older adults: Evidence from the China Health and Retirement Longitudinal Study. BMC Geriatr. 2022 Dec;22(1):537.

24. Hoogendijk EO, Smit AP, Van Dam C, Schuster NA, De Breij S, Holwerda TJ, et al. Frailty Combined with Loneliness or Social Isolation: An Elevated Risk for Mortality in Later Life. J Am Geriatr Soc. 2020 Nov;68(11):2587–93.

25. Hamaker EL, Kuiper RM, Grasman RPPP. A critique of the cross-lagged panel model. Psychol Methods. 2015;20(1):102–16.

26. Voelkle MC, Gische C, Driver CC, Lindenberger U. The Role of Time in the Quest for Understanding Psychological Mechanisms. Multivar Behav Res. 2018 Nov 2;53(6):782– 805.

27. Sliwinski MJ. Measurement[Burst Designs for Social Health Research. Soc Personal Psychol Compass. 2008 Jan;2(1):245–61.

28. Mitnitski A, Mogilner AJ, Rockwood K. Accumulation of Deficits as a Proxy Measure of Aging. Sci World J. 2001;1:323–36.

29. Theou O, Haviva C, Wallace L, Searle SD, Rockwood K. How to construct a frailty index from an existing dataset in 10 steps. Age Ageing. 2023 Dec 1;52(12):afad221.

30. Mayerl H, Stolz E, Freidl W. Frailty and depression: Reciprocal influences or common causes? Soc Sci Med. 2020 Oct;263:113273.

31. Stolz E, Mayerl H, Godin J, Hoogendijk EO, Theou O, Freidl W, et al. Reliability of the Frailty Index Among Community-Dwelling Older Adults. Lipsitz LA, editor. J Gerontol Ser A. 2023 Sep 20;glad227.

32. Hughes ME, Waite LJ, Hawkley LC, Cacioppo JT. A Short Scale for Measuring Loneliness in Large Surveys: Results From Two Population-Based Studies. Res Aging. 2004 Nov;26(6):655–72.

33. Curran PJ, Howard AL, Bainter SA, Lane ST, McGinley JS. The separation of between-person and within-person components of individual change over time: A latent curve model with structured residuals. J Consult Clin Psychol. 2014;82(5):879–94.

34. Usami S, Murayama K, Hamaker EL. A unified framework of longitudinal models to examine reciprocal relations. Psychol Methods. 2019 Oct;24(5):637–57.

35. Rohrer JM, Murayama K. These Are Not the Effects You Are Looking for: Causality and the Within-/Between-Persons Distinction in Longitudinal Data Analysis. Adv Methods Pract Psychol Sci. 2023 Jan;6(1):251524592211408.

36. Rioux C, Stickley ZL, Little TD. Solutions for latent growth modeling following COVID-19-related discontinuities in change and disruptions in longitudinal data collection. Int J Behav Dev. 2021 Sep;45(5):463–73.

37. Satorra A, Bentler PM. A scaled difference chi-square test statistic for moment structure analysis. Psychometrika. 2001 Dec;66(4):507–14.

38. Hu L, Bentler PM. Cutoff criteria for fit indexes in covariance structure analysis: Conventional criteria versus new alternatives. Struct Equ Model Multidiscip J. 1999 Jan;6(1):1–55.

39. R Core Team. R: A language and environment for statistical computing. [Internet]. Vienna, Austria: R Foundation for Statistical Computing; 2023. Available from: https://www.R-project.org/

40. Rosseel Y. lavaan: An R Package for Structural Equation Modeling. J Stat Softw. 2012;48(2):1–36.

41. Jorgensen TD, Pornprasertmanit S, Schoemann AM, Rosseel Y. semTools: Useful tools for structural equation modeling [Internet]. 2022. Available from: https://CRAN.R-project.org/package=semTools

42. Searle SD, Mitnitski A, Gahbauer EA, Gill TM, Rockwood K. A standard procedure for creating a frailty index. BMC Geriatr. 2008 Dec;8(1):24.

43. Christiansen J, Lund R, Qualter P, Andersen CM, Pedersen SS, Lasgaard M. Loneliness, Social Isolation, and Chronic Disease Outcomes. Ann Behav Med. 2021 Mar 20;55(3):203–15.

44. Utz RL, Swenson KL, Caserta M, Lund D, deVries B. Feeling Lonely Versus Being Alone: Loneliness and Social Support Among Recently Bereaved Persons. J Gerontol B Psychol Sci Soc Sci. 2014 Jan 1;69B(1):85–94.

45. Huxhold O, Henning G. The Risks of Experiencing Severe Loneliness Across Middle and Late Adulthood. Sneed R, editor. J Gerontol Ser B. 2023 Jul 11;78(10):1668–75.

46. Sheftel MG, Margolis R, Verdery AM. Life Events and Loneliness Transitions Among Middle-Aged and Older Adults Around the World. J Gerontol Ser B. 2024;gbad149.

47. Awad R, Shamay-Tsoory SG, Palgi Y. Fluctuations in loneliness due to changes in frequency of social interactions among older adults: a weekly based diary study. Int Psychogeriatr. 2023 Jun;35(6):293–303.

48. Mitnitski A, Song X, Rockwood K. Trajectories of changes over twelve years in the health status of Canadians from late middle age. Exp Gerontol. 2012 Dec;47(12):893–9.

49. Luo Y, Hawkley LC, Waite LJ, Cacioppo JT. Loneliness, health, and mortality in old age: A national longitudinal study. Soc Sci Med. 2012 Mar;74(6):907–14.

50. Luo Y, Waite LJ. Loneliness and Mortality Among Older Adults in China. J Gerontol B Psychol Sci Soc Sci. 2014 Jul 1;69(4):633–45.

51. Feenstra M, Oud FMM, Jansen CJ, Smidt N, Van Munster BC, De Rooij SE. Reproducibility and responsiveness of the Frailty Index and Frailty Phenotype in older hospitalized patients. BMC Geriatr. 2021 Dec;21(1):499.

52. Alsubheen SA, Oliveira A, Habash R, Goldstein R, Brooks D. Systematic review of psychometric properties and cross-cultural adaptation of the University of California and Los Angeles loneliness scale in adults. Curr Psychol. 2023 May;42(14):11819–33.

53. Maes M, Qualter P, Lodder GMA, Mund M. How (Not) to Measure Loneliness: A Review of the Eight Most Commonly Used Scales. Int J Environ Res Public Health. 2022 Aug 30;19(17):10816.

54. Courtin E, Knapp M. Social isolation, loneliness and health in old age: a scoping review. Health Soc Care Community. 2017 May;25(3):799–812.

55. Giné-Garriga M, Jerez-Roig J, Coll-Planas L, Skelton DA, Inzitari M, Booth J, et al. Is loneliness a predictor of the modern geriatric giants? Analysis from the survey of health, ageing, and retirement in Europe. Maturitas. 2021 Feb;144:93–101.

56. Ong AD, Uchino BN, Wethington E. Loneliness and Health in Older Adults: A Mini-Review and Synthesis. Gerontology. 2016;62(4):443–9.

57. Cacioppo JT, Cacioppo S, Capitanio JP, Cole SW. The Neuroendocrinology of Social Isolation. Annu Rev Psychol. 2015 Jan 3;66(1):733–67.

58. Bessa B, Ribeiro O, Coelho T. Assessing the social dimension of frailty in old age: A systematic review. Arch Gerontol Geriatr. 2018 Sep;78:101–13.

59. Mayerl H, Stolz E, Freidl W. Lonely and depressed in older age: prospective associations and common vulnerabilities. Aging Ment Health. 2023 Mar 4;27(3):640–5.

60. Buttery AK, Busch MA, Gaertner B, Scheidt-Nave C, Fuchs J. Prevalence and correlates of frailty among older adults: findings from the German health interview and examination survey. BMC Geriatr. 2015 Dec;15(1):22.

61. Kikusui T, Winslow JT, Mori Y. Social buffering: relief from stress and anxiety. Philos Trans R Soc B Biol Sci. 2006 Dec 29;361(1476):2215–28.

62. Olaya B, Domènech-Abella J, Moneta MV, Lara E, Caballero FF, Rico-Uribe LA, et al. All-cause mortality and multimorbidity in older adults: The role of social support and loneliness. Exp Gerontol. 2017 Dec;99:120–6.

63. Theou O, O‘Connell MDL, King-Kallimanis BL, O’Halloran AM, Rockwood K, Kenny RA. Measuring frailty using self-report and test-based health measures. Age Ageing. 2015 May;44(3):471–7.

64. Allison PD. Missing Data Techniques for Structural Equation Modeling. J Abnorm Psychol. 2003 Nov;112(4):545–57.

65. Stolz E. FRequent health Assessment In Later life (FRAIL70+) (SUF edition) [Internet]. AUSSDA; 2024. Available from: https://data.aussda.at/citation?persistentId=doi:10.11587/DJNOHX

