## Supplementary Material for "Frailty and loneliness among community-dwelling older adults: Examining reciprocal associations within a measurement burst design"

#### Contents

**Supplementary Methods 1: Data collection process**

**Supplementary Figure 1: Interview schedule**

**Supplementary Table 1: Health deficits considered in the FI**

**Supplementary Table 2: UCLA loneliness scale**

**Supplementary Methods 2: Model selection**

**Supplementary Figure 2: Correlation between FI and LS**

**Supplementary Table 3: Univariate FI models**

**Supplementary Table 4: Comparison of FI models**

**Supplementary Table 5: Univariate LS models**

**Supplementary Table 6: Comparison of LS models**

**Supplementary Table 7: Multiple group analysis with regard to sex**

**Supplementary Table 8: Multiple group analysis with regard to age**

**Supplementary Table 9: Multiple group analysis with regard to living alone**

**Supplementary Table 10: Multiple group analysis with regard to social participation**

**Supplementary Table 11: LCM-SR with bedrest as time varying covariate**

**Supplementary Table 12: LCM-SR with falls as time varying covariate**

**Supplementary References**

### Supplementary Methods 1: Data collection process

A professional survey agency contacted community-dwelling older adults based on previous participation in population representative surveys. Interviewers explained the study's topic, duration (i. e., two rounds of seven biweekly interviews spaced one year apart), and the information required, ensured the anonymity of all personal data, and obtained written consent from participants before participation.

The first interview of the fist burst, conducted in person (computer assisted personal interview; CAPI) by the end of August 2021, lasted a median of 23.9 minutes. The subsequent computer assisted telephone interviews (CATI; waves 2 to 7) lasted between 8.7 and 9.5 minutes (median; Stolz, 2024). With a total of 40 participants, all interviews were conducted in person due to continued physical performance tests (i. e., grip strength, gait speed, and chair rise; physical performance tests are not considered in the current analysis). The mean duration of the first burst was 87 days ($SD$=13), i. e., on average, the interviews were repeated at intervals of 14.5 days.

Interviews of the second burst (w8–w14) stared in November 2022 (i. e., one year after the end of the first burst; the average number of days between w7 and w8 was 376, $SD$=11). Again, the first interview of the second burst (w8) was conducted in person (CAPI) and lasted a median of 15.1 minutes. The subsequent interviews (w9–w14) were conducted via telephone (CATI) and lasted between 7.9 and 8.7 minutes (median; Stolz, 2024). The mean duration of the second burst was 76 days ($SD$=11), i. e., on average, the interviews were repeated at intervals of 12.7 days.

Supplementary Figure 1: Interview schedule in which individuals and completed interviews are depicted with gray lines and black circles, respectively

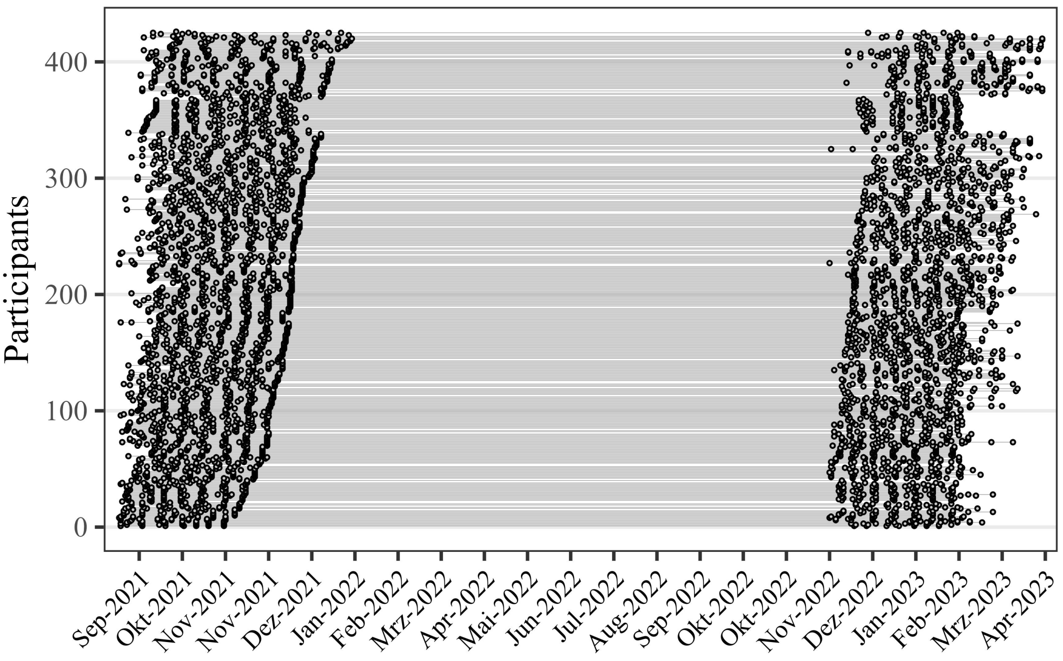

Supplementary Table 1: Health deficits considered in the frailty index

| | health deficit | assigned values | prevalence at baseline (%) | missing data at baseline (%) |
|---|---|---|---|---|
| 1 | Self-rated health | 0 = excellent<br>0.25 = very good<br>0.5 = good<br>0.75 = moderate<br>1 = poor | 0 = 6.3<br>0.25 = 18.5<br>0.5 = 35.9<br>0.75 = 28.4<br>1 = 10.8 | – |
| 2 | Dizziness | 0 = no/1 = yes | 1 = 20.7 | – |
| 3 | Pain (rating from 0–10) | 0 = 0<br>0.5 = $\geq$ 1 & $\leq$3<br>1 = $\geq$ | 0 = 26.1<br>0.5 = 35.2<br>1 = 38.7 | – |
| 4 | Tiredness | 0 = never<br>0.5 = sometimes<br>1 = often | 0 = 43.0<br>0.5 = 42.0<br>1 = 15.0 | – |
| 5 | Vision | 0 = excellent<br>0.25 = very good<br>0.5 = good<br>0.75 = moderate<br>1 = poor | 0 = 10.4<br>0.25 = 38.0<br>0.5 = 35.1<br>0.75 = 13.4<br>1 = 3.1 | 0.5 |
| 6 | Hearing | 0 = excellent<br>0.25 = very good<br>0.5 = good<br>0.75 = moderate<br>1 = poor | 0 = 12.7<br>0.25 = 36.2<br>0.5 = 30.8<br>0.75 = 16.9<br>1 = 3.3 | 0.2 |
| 7 | Attention (10 words immediate recall test) | 0 = $\geq$ 5 words<br>1 = $<$ 5 words | 1 = 20.0 | – |
| 8 | Memory (10 words delayed recall test) | 0 = $\geq$ 4 words<br>1 = $<$ 4 words | 1 = 32.6 | – |
| 9 | Physical inactivity (moderate physical activity) | 0 = every day/almost every day & multiple times a week<br>1 = once per week & less often | 1 = 21.4 | – |
| Doctor told you had: . . . | | | | |
| 10 | Heart problem (myocardial infarction, coronary thrombosis, other problem including congestive heart failure) | 0 = no/1 = yes | 1 = 15.3 | – |
| 11 | High blood pressure or hypertension | 0 = no/1 = yes | 1 = 48.6 | – |
| 12 | Stroke or cerebral vascular disease | 0 = no/1 = yes | 1 = 4.7 | – |
| 13 | Diabetes or high blood sugar | 0 = no/1 = yes | 1 = 19.5 | – |
| 14 | Chronic lung disease such as chronic bronchitis or emphysema | 0 = no/1 = yes | 1 = 9.9 | – |
| 15 | Cancer or malignant tumor, including leukemia or lymphoma | 0 = no/1 = yes | 1 = 5.6 | – |
| 16 | Arthritis, including osteoarthritis, or rheumatism | 0 = no/1 = yes | 1 = 27.0 | – |
| 17 | Chronic renal disease | 0 = no/1 = yes | 1 = 2.8 | – |
| 18 | Alzheimer's disease, dementia or any other serious memory impairment | 0 = no/1 = yes | 1 = 3.1 | – |
| 19 | Difficulty getting dressed | 0 = no/1 = yes | 1 = 12.2 | – |
| 20 | Difficulty walking across room | 0 = no/1 = yes | 1 = 8.0 | 0.5 |
| 21 | Difficulty bathing/showering | 0 = no/1 = yes | 1 = 9.9 | – |
| 22 | Difficulty eating | 0 = no/1 = yes | 1 = 3.8 | – |
| 23 | Difficulty going in/out of bed | 0 = no/1 = yes | 1 = 7.5 | – |
| 24 | Difficulty using toilet | 0 = no/1 = yes | 1 = 3.5 | – |
| 25 | Difficulty preparing a warm meal | 0 = no/1 = yes | 1 = 5.4 | 0.5 |
| 26 | Difficulty shopping groceries | 0 = no/1 = yes | 1 = 11.8 | 0.7 |
| 27 | Difficulty using telephone | 0 = no/1 = yes | 1 = 1.6 | – |
| 28 | Difficulty taking medicine | 0 = no/1 = yes | 1 = 1.9 | 1.6 |
| 29 | Difficulty walking 100 meters | 0 = no/1 = yes | 1 = 12.5 | 0.7 |
| 30 | Difficulty taking one flight of stairs | 0 = no/1 = yes | 1 = 23.6 | 0.7 |
| 31 | Difficulty reaching or extending your arms above shoulder level | 0 = no/1 = yes | 1 = 14.8 | – |
| 32 | Difficulty lifting or carrying weights over 10 pounds/5 kilos, like a heavy bag of groceries | 0 = no/1 = yes | 1 = 27.8 | 0.5 |
| 33 | Difficulty concentrating | 0 = never/rarely<br>0.5 = sometimes<br>1 = often/always | 0 = 71.4<br>0.5 = 25.8<br>1 = 2.8 | – |
| 34 | Everything takes effort | 0 = never/rarely<br>0.5 = sometimes<br>1 = often/always | 0 = 66.9<br>0.5 = 23.9<br>1 = 9.2 | – |
| 35 | Sleep problems | 0 = never/rarely<br>0.5 = sometimes<br>1 = often/always | 0 = 52.7<br>0.5 = 35.3<br>1 = 12.0 | 0.2 |
| 36 | Could not get going | 0 = never/rarely<br>0.5 = sometimes<br>1 = often/always | 0 = 65.7<br>0.5 =27.7<br>1 = 6.6 | – |
| 37 | Poor appetite | 0 = never/rarely<br>0.5 = sometimes<br>1 = often/always | 0 = 89.0<br>0.5 = 8.0<br>1 = 3.1 | – |

Supplementary Table 2: University of California, Los Angeles – Loneliness Scale

| **Please tell me how often the following applied to you in the last two weeks:** | | rarely or never | sometimes | often or always | don't know/can't say |
|---|---|---|---|---|---|
| 1 | I felt like I was lacking companionship | (1) | (2) | (3) | (999) |
| 2 | I felt left out | (1) | (2) | (3) | (999) |
| 3 | I felt isolated from others | (1) | (2) | (3) | (999) |

#### Supplementary Methods 2: Model selection

To study individual change over time, two frameworks exist: the mixed-effects (ME) approach and the latent-curve (LC) approach. While previous methodological research has shown that these two modeling frameworks share some overlap and estimates align with optimal data (see for instance McNeish and Matta, 2018 for a brief overview).

Since our analytical focus lies on the decomposition of within- and between-person effects in short-term dynamics (i.e., over weeks and months) between frailty and loneliness, a “complex” residual structure is necessary. In other words, we aim to examine the 1) separate autocorrelation of frailty and loneliness, 2) (within-person) cross-lagged effect of frailty on loneliness (i. e., frailty levels that are higher/lower than usual for a person at time t1 predict corresponding levels of loneliness that are higher/lower than usual at time t2) and loneliness on frailty, and 3) (within-person) within-time relationship between frailty and loneliness (i. e., frailty levels that are higher/lower than usual for a person at time t1 are associated with loneliness levels that are higher/lower than usual at time t1). While both ME and LC frameworks allow the specification of residual structures, the ME framework is often constrained by preprogrammed software options, whereas the LC framework allows the definition of any structure (McNeish & Matta, 2018). Therefore, we used a latent curve model with structured residuals (LCM-SR; Curran et al., 2014) to separate between-person variability from within-person variability.

Supplementary Figure 2: Pairwise correlations

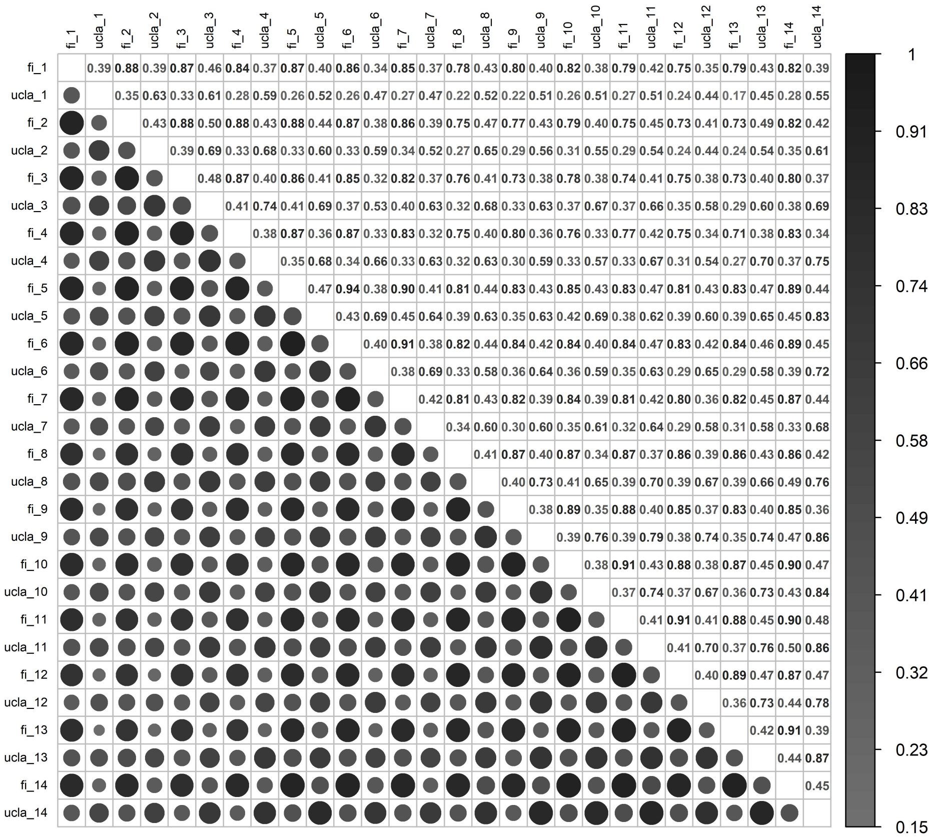

Supplementary Table 3: Parameters of FI models 1, 2, and 3

| | model 1 | model 2 | model 3 |
|---|---|---|---|
| Parameter | Est.[95%CI] | Est.[95%CI] | Est.[95%CI] |
| Random effects: Means | | | |
| Intercept FI1* | 0.18 [0.17, 0.19] | 0.18 [0.17, 0.19] | 0.18 [0.17, 0.19] |
| Intercept FI2* | 0.20 [0.19, 0.21] | 0.20 [0.19, 0.22] | 0.20 [0.19, 0.22] |
| Slope FI1* | – | – | –0.00 [–0.00, 0.00] |
| Slope FI2* | – | – | 0.00 [–0.00, 0.00] |
| Random Effects: Correlation ($\zeta$) | | | |
| Intercept FI1 $\leftrightarrow$ Intercept FI2 | – | 0.94 [0.92, 0.97] | 0.95 [0.93, 0.97] |
| Autoregressive (FI $\rightarrow$ FI) | | | |
| $\alpha_1$ | 0.89 [0.86, 0.92] | 0.26 [0.12, 0.40] | 0.26 [0.12, 0.40] |
| $\alpha_2$ | 0.89 [0.84, 0.93] | 0.31 [0.17, 0.45] | 0.31 [0.17, 0.45] |
| $\alpha_3$ | 0.87 [0.82, 0.93] | 0.29 [0.05, 0.54] | 0.29 [0.05, 0.54] |
| $\alpha_4$ | 0.88 [0.83, 0.93] | 0.08 [–0.10, 0.25] | 0.08 [–0.10, 0.25] |
| $\alpha_5$ | 0.94 [0.93, 0.96] | 0.27 [0.09, 0.45] | 0.27 [0.09, 0.45] |
| $\alpha_6$ | 0.92 [0.89, 0.94] | 0.20 [0.02, 0.37] | 0.20 [0.02, 0.37] |
| $\alpha_7$ | 0.84 [0.80, 0.88] | 0.14 [–0.12, 0.39] | 0.14 [–0.12, 0.39] |
| $\alpha_8$ | 0.89 [0.85, 0.93] | 0.22 [0.06, 0.38] | 0.22 [0.06, 0.38] |
| $\alpha_9$ | 0.91 [0.87, 0.94] | 0.17 [–0.02, 0.37] | 0.17 [–0.03, 0.37] |
| $\alpha_{10}$ | 0.92 [0.90, 0.95] | 0.10 [–0.23, 0.44] | 0.10 [–0.24, 0.44] |
| $\alpha_{11}$ | 0.93 [0.90, 0.96] | 0.29 [0.07, 0.52] | 0.29 [0.07, 0.51] |
| $\alpha_{12}$ | 0.90 [0.85, 0.95] | 0.22 [0.03, 0.41] | 0.22 [0.03, 0.41] |
| $\alpha_{13}$ | 0.92 [0.89, 0.96] | 0.35 [0.10, 0.60] | 0.35 [0.11, 0.60] |

*Note.* FI = Frailty Index. In model 1 only fixed intercepts are specified. For models 2 and 3 random intercepts and fixed slopes are added, respectively. We report standardized parameter estimates and 95%-CI for all variables, except for variables with * in superscript. Here, unstandardized estimates are reported instead.

Supplementary Table 4: Model fit statistics and comparison of FI-models 1, 2, and 3

| model | $\chi^2(df)$ | $p-value$ | ref. | $\Delta_{\chi^2}(\Delta df)$ | $\Delta p-value$ | TLI | CFI | SRMR | RMSEA [90% CI] | AIC | BIC |
|---|---|---|---|---|---|---|---|---|---|---|---|
| 1 | 1227.65(90) | <.001 | – | – | – | 0.873 | 0.874 | 0.268 | 0.190 [0.177, 0.204] | –13192 | –13074 |
| 2 | 230.76(87) | <.001 | 1 | 109.07(3) | <.001 | 0.983 | 0.983 | 0.034 | 0.070 [0.055, 0.086] | –14183 | –14053 |
| 3 | 230.60(85) | <.001 | 2 | 0.076(2) | 0.931 | 0.982 | 0.983 | 0.034 | 0.072 [0.056, 0.087] | –14179 | –14041 |

*Note.* $\chi^2$ = model fit statistic; $df$ = degrees of freedom, ref. = reference model; $\Delta_{\chi^2}$ = Satorra-Bentler scaled chi-square difference test; $\Delta df$ = differences in degrees of freedom; TLI = robust Tucker-Lewis-index; CFI = robust comparative-fit-index; SRMR = standardized root mean square residual; RMSEA = robust root mean square error of approximation; CI = confidence interval; AIC = Akaike information criterion; BIC = sample-size adjusted Bayesian information criterion.

#### Supplementary Table 5: Parameters of LS-models 1, 2, 3, and 4

| | model 1 | model 2 | model 3 | model 4 |
|---|---|---|---|---|
| Parameter | Est.[95%CI] | Est.[95%CI] | Est.[95%CI] | Est.[95%CI] |
| Random effects: Means | | | | |
| Intercept LS1* | 3.46 [3.37, 3.54] | 3.45 [3.36, 3.53] | 3.39 [3.30, 3.48] | 3.39 [3.30, 3.49] |
| Intercept LS2* | 3.41 [3.33, 3.50] | 3.48 [3.39, 3.58] | 3.51 [3.41, 3.61] | 3.51 [3.41, 3.61] |
| Slope LS1 | – | – | 0.02 [0.00, 0.03] | 0.02 [0.00, 0.03] |
| Slope LS2 | – | – | –0.01 [–0.02, 0.00] | –0.01 [–0.02, 0.00] |
| Random Effects: Correlation ($\zeta$) | | | | |
| Intercept LS1 ↔ Intercept LS2 | – | 0.92 [0.86, 0.98] | 0.92 [0.86, 0.98] | 0.83 [0.71, 0.96] |
| Intercept LS1 ↔ Slope LS1 | – | – | – | –0.14 [–0.55, 0.27] |
| Intercept LS2 ↔ Slope LS1 | – | – | – | 0.28 [–0.16, 0.72] |
| Autoregressive (LS → LS) | | | | |
| $\beta_1$ | 0.64 [0.52, 0.77] | 0.25 [0.09, 0.42] | 0.26 [0.09, 0.42] | 0.13 [–0.13, 0.38] |
| $\beta_2$ | 0.70 [0.58, 0.82] | 0.20 [–0.03, 0.43] | 0.19 [–0.04, 0.42] | 0.09 [–0.19, 0.36] |
| $\beta_3$ | 0.75 [0.62, 0.88] | 0.18 [–0.11, 0.46] | 0.17 [–0.12, 0.45] | 0.13 [–0.11, 0.38] |
| $\beta_4$ | 0.71 [0.59, 0.83] | 0.01 [–0.22, 0.23] | 0.01 [–0.22, 0.24] | 0.02 [–0.22, 0.26] |
| $\beta_5$ | 0.69 [0.57, 0.81] | 0.15 [–0.08, 0.39] | 0.15 [–0.09, 0.38] | 0.12 [–0.20, 0.43] |
| $\beta_6$ | 0.69 [0.60, 0.78] | 0.27 [0.09, 0.44] | 0.25 [0.08, 0.43] | 0.18 [–0.07, 0.42] |
| $\beta_7$ | 0.61 [0.49, 0.72] | 0.01 [–0.25, 0.27] | –0.01 [–0.27, 0.25] | 0.01 [–0.27, 0.29] |
| $\beta_8$ | 0.74 [0.64, 0.84] | 0.08 [–0.16, 0.31] | 0.08 [–0.16, 0.31] | 0.08 [–0.15, 0.31] |
| $\beta_9$ | 0.77 [0.66, 0.87] | 0.06 [–0.24, 0.36] | 0.06 [–0.23, 0.36] | 0.07 [–0.22, 0.35] |
| $\beta_{10}$ | 0.75 [0.64, 0.85] | 0.04 [–0.23, 0.30] | 0.03 [–0.23, 0.30] | 0.03 [–0.24, 0.30] |
| $\beta_{11}$ | 0.74 [0.64, 0.84] | –0.12 [–0.39, 0.15] | –0.12 [–0.39, 0.15] | –0.12 [–0.39, 0.15] |
| $\beta_{12}$ | 0.76 [0.66, 0.85] | 0.05 [–0.19, 0.28] | 0.04 [–0.20, 0.28] | 0.04 [–0.20, 0.27] |
| $\beta_{13}$ | 0.86 [0.78, 0.94] | 0.35 [0.09, 0.62] | 0.35 [0.08, 0.62] | 0.35 [0.08, 0.62] |

*Note.* LS = loneliness. In model 1 only fixed intercepts are specified. For models 2 and 3 random intercepts and fixed slopes are added, respectively. In model 4, a random and a fixed slope are specified. We report standardized parameter estimates and 95%-CI for all variables, except for variables with * in superscript. Here, unstandardized estimates are reported instead.

#### Supplementary Table 6: Model fit statistics and comparison of LS models 1, 2, 3, and 4

| model | | $\chi^2(df)$ | $p-value$ | ref. | $\Delta_{\chi^2}(\Delta df)$ | $\Delta p-value$ | TLI | CFI | SRMR | RMSEA [90% CI] | AIC | BIC |
|---|---|---|---|---|---|---|---|---|---|---|---|---|
| 1 | 1 | 1252.11(90) | <.001 | – | – | – | 0.763 | 0.766 | 0.383 | 0.187 [0.169, 0.205] | 11231 | 11348 |
| 2 | 2 | 297.02(87) | <.001 | 1 | 443.67(3) | <.001 | 0.987 | 0.988 | 0.060 | 0.043 [0.000, 0.072] | 10282 | 10411 |
| 3 | 3 | 287.94(85) | <.001 | 2 | 9.58 (2) | .008 | 0.988 | 0.989 | 0.059 | 0.041 [0.000, 0.071] | 10276 | 10414 |
| 4 | 4 | 273.03(82) | <.001 | 3 | 4 (3) | .261 | 0.989 | 0.990 | 0.049 | 0.040 [0.000, 0.071] | 10268 | 10418 |

*Note.* $\chi^2$ = model fit statistic; $df$ = degrees of freedom, ref. = reference model; $\Delta_{\chi^2}$ = Satorra-Bentler scaled chi-square difference test; $\Delta df$ = differences in degrees of freedom; TLI = robust Tucker-Lewis-index; CFI = robust comparative-fit-index; SRMR = standardized root mean square residual; RMSEA = robust root mean square error of approximation; CI = confidence interval; AIC = Akaike information criterion; BIC = sample-size adjusted Bayesian information criterion.

Supplementary Table 7: Multiple group analysis: Sex (female vs. male)

| | Female $n = 275$ | Male $n = 151$ |
|---|---|---|
| Parameter | Est.[95%CI] | Est.[95%CI] |
| Random effects: Means | | |
| Intercept FI1* | 0.20 [0.18, 0.21] | 0.15 [0.14, 0.17] |
| Intercept FI2* | 0.22 [0.20, 0.24] | 0.17 [0.15, 0.19] |
| Intercept LS1* | 3.46 [3.34, 3.58] | 3.27 [3.15, 3.38] |
| Intercept LS2* | 3.59 [3.45, 3.73] | 3.35 [3.24, 3.46] |
| Fixed effects: Means | | |
| Slope LS1* | 0.02 [0.00, 0.04] | 0.01 [−0.01, 0.03] |
| Slope LS2* | −0.01 [−0.03, 0.00] | −0.00 [−0.02, 0.01] |
| Random Effects: Correlation | | |
| $\zeta_1$: Intercept FI1 ↔ Intercept FI2 | 0.94 [0.90, 0.97] | 0.96 [0.93, 0.98] |
| $\zeta_2$: Intercept FI1 ↔ Intercept LS1 | 0.54 [0.40, 0.67] | 0.65 [0.49, 0.82] |
| $\zeta_3$: Intercept FI1 ↔ Intercept LS2 | 0.56 [0.42, 0.69] | 0.64 [0.48, 0.81] |
| $\zeta_4$: Intercept FI2 ↔ Intercept LS1 | 0.49 [0.35, 0.64] | 0.57 [0.41, 0.72] |
| $\zeta_5$: Intercept FI2 ↔ Intercept LS2 | 0.50 [0.34, 0.65] | 0.60 [0.46, 0.74] |
| $\zeta_6$: Intercept LS1 ↔ Intercept LS2 | 0.91 [0.83, 0.98] | 0.95 [0.86, 1.03] |
| Autoregressive (FI → FI) | | |
| $\alpha_1$ | 0.30 [0.11, 0.50] | 0.16 [−0.08, 0.41] |
| $\alpha_2$ | 0.30 [0.09, 0.50] | 0.30 [0.14, 0.46] |
| $\alpha_3$ | 0.35 [0.03, 0.66] | 0.15 [−0.16, 0.47] |
| $\alpha_4$ | 0.07 [−0.14, 0.29] | 0.05 [−0.35, 0.45] |
| $\alpha_5$ | 0.31 [0.10, 0.52] | 0.19 [−0.22, 0.60] |
| $\alpha_6$ | 0.27 [0.08, 0.46] | 0.04 [−0.32, 0.40] |
| $\alpha_7$ | 0.21 [−0.08, 0.49] | −0.19 [−0.47, 0.10] |
| $\alpha_8$ | 0.26 [0.10, 0.43] | 0.00 [−0.38, 0.38] |
| $\alpha_9$ | 0.29 [0.09, 0.50] | −0.17 [−0.63, 0.29] |
| $\alpha_{10}$ | −0.13 [−0.41, 0.14] | 0.46 [−0.06, 0.99] |
| $\alpha_{11}$ | 0.25 [−0.02, 0.51] | 0.38 [0.07, 0.68] |
| $\alpha_{12}$ | 0.22 [−0.05, 0.50] | 0.33 [0.02, 0.64] |
| $\alpha_{13}$ | 0.33 [0.06, 0.59] | 0.46 [0.04, 0.87] |
| Autoregressive (LS → LS) | | |
| $\beta_1$ | 0.23 [0.05, 0.42] | 0.29 [0.05, 0.53] |
| $\beta_2$ | 0.18 [−0.09, 0.45] | 0.12 [−0.18, 0.42] |
| $\beta_3$ | 0.07 [−0.25, 0.40] | 0.24 [−0.15, 0.63] |
| $\beta_4$ | 0.02 [−0.34, 0.38] | −0.13 [−0.54, 0.27] |
| $\beta_5$ | 0.11 [−0.22, 0.44] | 0.15 [−0.19, 0.50] |
| $\beta_6$ | 0.24 [0.03, 0.44] | 0.37 [0.02, 0.72] |
| $\beta_7$ | 0.08 [−0.22, 0.38] | −0.44 [−0.74, −0.14] |
| $\beta_8$ | 0.18 [−0.08, 0.43] | −0.24 [−0.60, 0.13] |
| $\beta_9$ | −0.04 [−0.35, 0.28] | 0.14 [−0.25, 0.54] |
| $\beta_{10}$ | −0.03 [−0.31, 0.26] | 0.04 [−0.43, 0.52] |
| $\beta_{11}$ | −0.09 [−0.44, 0.25] | −0.16 [−0.42, 0.10] |
| $\beta_{12}$ | −0.02 [−0.29, 0.25] | 0.11 [−0.35, 0.57] |
| $\beta_{13}$ | 0.18 [−0.25, 0.61] | 0.43 [0.15, 0.71] |
| Cross-lagged (LS → FI) | | |
| $\delta_1$ | 0.02 [−0.16, 0.21] | −0.03 [−0.32, 0.25] |
| $\delta_2$ | 0.08 [−0.08, 0.24] | 0.14 [−0.03, 0.31] |
| $\delta_3$ | 0.07 [−0.11, 0.24] | −0.00 [−0.25, 0.25] |
| $\delta_4$ | 0.11 [−0.15, 0.37] | −0.31 [−0.51, −0.12] |
| $\delta_5$ | −0.01 [−0.21, 0.18] | −0.15 [−0.40, 0.10] |
| $\delta_6$ | −0.01 [−0.15, 0.14] | −0.08 [−0.34, 0.18] |
| $\delta_7$ | 0.08 [−0.07, 0.22] | 0.10 [−0.19, 0.39] |
| $\delta_8$ | 0.05 [−0.16, 0.26] | −0.17 [−0.43, 0.09] |
| $\delta_9$ | 0.09 [−0.16, 0.34] | 0.04 [−0.24, 0.32] |
| $\delta_{10}$ | −0.13 [−0.40, 0.13] | 0.05 [−0.15, 0.26] |
| $\delta_{11}$ | 0.13 [−0.11, 0.36] | 0.06 [−0.20, 0.31] |
| $\delta_{12}$ | 0.07 [−0.18, 0.31] | −0.10 [−0.32, 0.11] |
| $\delta_{13}$ | 0.02 [−0.22, 0.26] | 0.04 [−0.11, 0.18] |
| Cross-lagged (FI → LS) | | |
| $\gamma_1$ | 0.09 [−0.04, 0.22] | 0.18 [0.02, 0.33] |
| $\gamma_2$ | 0.18 [0.01, 0.35] | 0.03 [−0.31, 0.38] |
| $\gamma_3$ | 0.15 [−0.09, 0.40] | 0.27 [0.11, 0.44] |
| $\gamma_4$ | 0.03 [−0.26, 0.32] | −0.26 [−0.47, −0.06] |
| $\gamma_5$ | −0.00 [−0.23, 0.23] | −0.05 [−0.36, 0.25] |
| $\gamma_6$ | −0.09 [−0.23, 0.05] | −0.09 [−0.32, 0.14] |
| $\gamma_7$ | 0.08 [−0.07, 0.23] | −0.11 [−0.33, 0.11] |
| $\gamma_8$ | 0.15 [−0.05, 0.34] | 0.18 [−0.25, 0.62] |
| $\gamma_9$ | −0.06 [−0.30, 0.18] | 0.23 [−0.09, 0.54] |
| $\gamma_{10}$ | 0.18 [−0.08, 0.44] | −0.02 [−0.36, 0.32] |
| $\gamma_{11}$ | 0.20 [−0.08, 0.47] | 0.22 [0.01, 0.44] |
| $\gamma_{12}$ | −0.12 [−0.33, 0.10] | 0.19 [−0.23, 0.62] |
| $\gamma_{13}$ | 0.34 [0.03, 0.65] | 0.03 [−0.09, 0.15] |
| Within-time (FI ↔ LS) | | |
| $\lambda_1$ | 0.21 [0.08, 0.34] | 0.31 [0.10, 0.52] |
| $\lambda_2$ | 0.21 [0.08, 0.34] | 0.02 [−0.25, 0.29] |
| $\lambda_3$ | 0.19 [0.06, 0.33] | 0.15 [−0.02, 0.32] |
| $\lambda_4$ | 0.22 [0.01, 0.42] | 0.08 [−0.11, 0.26] |
| $\lambda_5$ | 0.20 [−0.09, 0.50] | 0.00 [−0.31, 0.32] |
| $\lambda_6$ | 0.15 [−0.01, 0.32] | 0.30 [0.02, 0.57] |
| $\lambda_7$ | 0.22 [0.08, 0.37] | 0.03 [−0.15, 0.21] |
| $\lambda_8$ | 0.10 [−0.07, 0.26] | −0.01 [−0.38, 0.36] |
| $\lambda_9$ | 0.11 [−0.10, 0.31] | −0.01 [−0.40, 0.39] |
| $\lambda_{10}$ | 0.06 [−0.18, 0.31] | 0.23 [−0.02, 0.49] |
| $\lambda_{11}$ | 0.12 [−0.15, 0.40] | 0.14 [−0.03, 0.30] |
| $\lambda_{12}$ | −0.13 [−0.30, 0.05] | 0.46 [0.04, 0.87] |
| $\lambda_{13}$ | 0.12 [−0.06, 0.31] | −0.02 [−0.17, 0.13] |
| $\lambda_{14}$ | 0.11 [−0.09, 0.32] | 0.10 [−0.12, 0.31] |

*Note.* We report standardized parameter estimates and 95.00 % CI intervals for all variables, except for the means of fixed/random effects; here, we report unstandardized estimates, as indicated by *.
Model fit: $\chi^2(648) = 1461.66$ $p < .001$; robust TLI = 0.928; robust CFI = 0.938; SRMR = 0.069; robust RMSEA [90% CI] = 0.088 [0.077, 0.098].

#### Supplementary Table 8: Multiple group analysis: Age ($\leq$ 74 vs. 75–79 vs. $\geq$ 80)

| | $\leq$ 74<br>$n$ = 166 | 75–79<br>$n$ = 127 | $\geq$ 80<br>$n$ = 133 |
|---|---|---|---|
| Parameter | Est.[95%CI] | Est.[95%CI] | Est.[95%CI] |
| Random effects: Means | | | |
| Intercept FI1* | 0.14 [0.12, 0.15] | 0.18 [0.16, 0.20] | 0.24 [0.21, 0.27] |
| Intercept FI2* | 0.15 [0.13, 0.17] | 0.21 [0.18, 0.23] | 0.27 [0.24, 0.29] |
| Intercept LS1* | 3.30 [3.19, 3.41] | 3.35 [3.17, 3.52] | 3.57 [3.37, 3.76] |
| Intercept LS2* | 3.31 [3.20, 3.42] | 3.40 [3.26, 3.54] | 3.82 [3.58, 4.05] |
| Fixed effects: Means | | | |
| Slope LS1* | 0.01 [−0.01, 0.02] | 0.02 [−0.01, 0.05] | 0.03 [−0.00, 0.06] |
| Slope LS2* | 0.00 [−0.01, 0.02] | 0.00 [−0.02, 0.03] | −0.02 [−0.04, 0.00] |
| Random Effects: Correlation | | | |
| $\zeta_1$: Intercept FI1 ↔ Intercept FI2 | 0.91 [0.86, 0.96] | 0.95 [0.91, 0.98] | 0.95 [0.91, 0.99] |
| $\zeta_2$: Intercept FI1 ↔ Intercept LS1 | 0.59 [0.45, 0.73] | 0.66 [0.47, 0.86] | 0.49 [0.30, 0.68] |
| $\zeta_3$: Intercept FI1 ↔ Intercept LS2 | 0.54 [0.38, 0.69] | 0.54 [0.30, 0.79] | 0.55 [0.37, 0.74] |
| $\zeta_4$: Intercept FI2 ↔ Intercept LS1 | 0.48 [0.32, 0.65] | 0.58 [0.41, 0.75] | 0.45 [0.25, 0.66] |
| $\zeta_5$: Intercept FI2 ↔ Intercept LS2 | 0.45 [0.27, 0.63] | 0.50 [0.28, 0.71] | 0.53 [0.32, 0.73] |
| $\zeta_6$: Intercept LS1 ↔ Intercept LS2 | 0.93 [0.87, 0.99] | 0.90 [0.73, 1.07] | 0.94 [0.86, 1.01] |
| Autoregressive (FI → FI) | | | |
| $\alpha_1$ | 0.31 [0.08, 0.54] | 0.11 [−0.23, 0.44] | 0.35 [0.08, 0.62] |
| $\alpha_2$ | 0.32 [0.14, 0.50] | 0.21 [−0.07, 0.50] | 0.39 [0.06, 0.71] |
| $\alpha_3$ | 0.40 [−0.04, 0.85] | 0.37 [0.00, 0.73] | 0.29 [−0.12, 0.70] |
| $\alpha_4$ | 0.01 [−0.52, 0.54] | 0.30 [0.02, 0.58] | 0.11 [−0.23, 0.45] |
| $\alpha_5$ | 0.28 [0.00, 0.55] | 0.29 [0.01, 0.58] | 0.33 [0.02, 0.65] |
| $\alpha_6$ | 0.06 [−0.25, 0.36] | 0.26 [−0.02, 0.54] | 0.24 [−0.04, 0.52] |
| $\alpha_7$ | −0.04 [−0.31, 0.23] | 0.35 [−0.18, 0.88] | 0.00 [−0.34, 0.35] |
| $\alpha_8$ | 0.24 [0.01, 0.47] | 0.08 [−0.23, 0.40] | 0.25 [−0.03, 0.54] |
| $\alpha_9$ | 0.36 [0.16, 0.56] | 0.15 [−0.24, 0.53] | −0.05 [−0.43, 0.34] |
| $\alpha_{10}$ | −0.27 [−0.62, 0.08] | 0.47 [−0.00, 0.93] | −0.10 [−0.57, 0.36] |
| $\alpha_{11}$ | 0.34 [−0.00, 0.67] | 0.18 [−0.20, 0.56] | 0.44 [0.12, 0.75] |
| $\alpha_{12}$ | 0.24 [−0.25, 0.73] | 0.17 [−0.10, 0.43] | 0.18 [−0.10, 0.47] |
| $\alpha_{13}$ | 0.25 [−0.06, 0.56] | 0.35 [−0.03, 0.74] | 0.50 [0.18, 0.83] |
| Autoregressive (LS → LS) | | | |
| $\beta_1$ | 0.07 [−0.36, 0.49] | 0.13 [−0.05, 0.30] | 0.49 [0.27, 0.70] |
| $\beta_2$ | 0.00 [−0.27, 0.27] | −0.18 [−0.56, 0.19] | 0.53 [0.25, 0.80] |
| $\beta_3$ | 0.35 [0.07, 0.64] | −0.14 [−0.71, 0.43] | 0.25 [−0.02, 0.52] |
| $\beta_4$ | 0.09 [−0.21, 0.38] | 0.09 [−0.29, 0.47] | 0.07 [−0.31, 0.44] |
| $\beta_5$ | 0.02 [−0.26, 0.30] | 0.13 [−0.23, 0.50] | 0.29 [−0.08, 0.65] |
| $\beta_6$ | 0.29 [−0.02, 0.60] | 0.37 [0.02, 0.72] | 0.23 [−0.07, 0.54] |
| $\beta_7$ | 0.21 [−0.32, 0.73] | −0.19 [−0.54, 0.17] | 0.05 [−0.31, 0.41] |
| $\beta_8$ | −0.02 [−0.56, 0.52] | 0.06 [−0.38, 0.50] | −0.03 [−0.31, 0.25] |
| $\beta_9$ | −0.44 [−0.94, 0.06] | −0.20 [−0.73, 0.34] | 0.37 [−0.05, 0.79] |
| $\beta_{10}$ | 0.15 [−0.13, 0.43] | 0.00 [−0.62, 0.62] | 0.00 [−0.44, 0.44] |
| $\beta_{11}$ | 0.10 [−0.33, 0.53] | −0.23 [−0.68, 0.23] | −0.07 [−0.60, 0.46] |
| $\beta_{12}$ | 0.08 [−0.49, 0.64] | −0.08 [−0.38, 0.22] | 0.27 [−0.17, 0.71] |
| $\beta_{13}$ | 0.36 [0.14, 0.58] | 0.32 [−0.36, 1.00] | 0.47 [−0.07, 1.00] |
| Cross-lagged (LS → FI) | | | |
| $\delta_1$ | −0.05 [−0.31, 0.21] | 0.05 [−0.26, 0.36] | −0.02 [−0.23, 0.18] |
| $\delta_2$ | −0.03 [−0.21, 0.14] | 0.19 [−0.08, 0.45] | 0.21 [0.04, 0.38] |
| $\delta_3$ | −0.04 [−0.30, 0.22] | 0.09 [−0.26, 0.43] | −0.01 [−0.24, 0.22] |
| $\delta_4$ | −0.05 [−0.35, 0.26] | −0.12 [−0.36, 0.12] | 0.14 [−0.17, 0.44] |
| $\delta_5$ | 0.11 [−0.07, 0.28] | −0.11 [−0.41, 0.19] | −0.17 [−0.43, 0.08] |
| $\delta_6$ | 0.14 [−0.09, 0.36] | −0.03 [−0.33, 0.27] | −0.12 [−0.31, 0.08] |
| $\delta_7$ | 0.07 [−0.08, 0.23] | 0.01 [−0.16, 0.18] | 0.11 [−0.16, 0.38] |
| $\delta_8$ | −0.06 [−0.19, 0.07] | 0.17 [−0.13, 0.48] | −0.10 [−0.41, 0.20] |
| $\delta_9$ | −0.10 [−0.29, 0.09] | −0.19 [−0.45, 0.07] | 0.25 [−0.16, 0.66] |
| $\delta_{10}$ | −0.09 [−0.37, 0.18] | −0.04 [−0.23, 0.15] | −0.07 [−0.81, 0.67] |
| $\delta_{11}$ | 0.01 [−0.19, 0.22] | 0.29 [−0.01, 0.60] | −0.12 [−0.37, 0.14] |
| $\delta_{12}$ | 0.27 [−0.06, 0.61] | −0.33 [−0.60, −0.07] | 0.14 [−0.14, 0.42] |
| $\delta_{13}$ | 0.23 [0.06, 0.40] | 0.18 [−0.12, 0.48] | −0.08 [−0.45, 0.28] |
| Cross-lagged (FI → LS) | | | |
| $\gamma_1$ | 0.15 [−0.02, 0.32] | 0.13 [−0.13, 0.39] | −0.00 [−0.14, 0.13] |
| $\gamma_2$ | 0.20 [−0.07, 0.46] | 0.38 [−0.02, 0.77] | 0.04 [−0.17, 0.26] |
| $\gamma_3$ | 0.20 [−0.09, 0.49] | 0.07 [−0.21, 0.36] | 0.33 [0.01, 0.66] |
| $\gamma_4$ | −0.06 [−0.25, 0.14] | 0.16 [−0.21, 0.53] | −0.30 [−0.61, 0.01] |
| $\gamma_5$ | 0.09 [−0.22, 0.40] | −0.22 [−0.60, 0.16] | 0.13 [−0.19, 0.45] |
| $\gamma_6$ | −0.09 [−0.35, 0.17] | −0.01 [−0.25, 0.24] | −0.27 [−0.46, -0.08] |
| $\gamma_7$ | −0.06 [−0.23, 0.11] | 0.14 [−0.06, 0.33] | 0.04 [−0.24, 0.32] |
| $\gamma_8$ | 0.04 [−0.14, 0.22] | 0.01 [−0.26, 0.29] | 0.17 [−0.18, 0.51] |
| $\gamma_9$ | −0.05 [−0.36, 0.25] | −0.11 [−0.42, 0.21] | −0.00 [−0.42, 0.42] |
| $\gamma_{10}$ | −0.09 [−0.32, 0.14] | 0.17 [−0.10, 0.44] | 0.01 [−0.57, 0.58] |
| $\gamma_{11}$ | 0.24 [−0.09, 0.56] | −0.04 [−0.31, 0.22] | 0.36 [−0.10, 0.82] |
| $\gamma_{12}$ | 0.22 [−0.08, 0.51] | 0.08 [−0.25, 0.42] | −0.12 [−0.43, 0.20] |
| $\gamma_{13}$ | 0.55 [0.24, 0.86] | 0.08 [−0.48, 0.65] | 0.15 [−0.12, 0.43] |
| Within-time (FI ↔ LS) | | | |
| $\lambda_1$ | 0.06 [−0.17, 0.29] | 0.25 [0.07, 0.43] | 0.38 [0.20, 0.57] |
| $\lambda_2$ | 0.08 [−0.16, 0.31] | 0.34 [0.14, 0.53] | 0.10 [−0.07, 0.26] |
| $\lambda_3$ | 0.20 [0.03, 0.37] | 0.24 [−0.04, 0.52] | 0.17 [−0.02, 0.35] |
| $\lambda_4$ | −0.09 [−0.30, 0.13] | 0.09 [−0.16, 0.34] | 0.21 [−0.04, 0.46] |
| $\lambda_5$ | 0.20 [−0.04, 0.43] | 0.14 [−0.25, 0.54] | 0.13 [−0.14, 0.41] |
| $\lambda_6$ | 0.26 [0.01, 0.51] | 0.03 [−0.37, 0.44] | 0.24 [−0.06, 0.54] |
| $\lambda_7$ | 0.04 [−0.11, 0.20] | 0.19 [−0.06, 0.44] | 0.22 [0.02, 0.42] |
| $\lambda_8$ | 0.16 [−0.01, 0.33] | 0.09 [−0.13, 0.32] | −0.01 [−0.27, 0.25] |
| $\lambda_9$ | −0.15 [−0.33, 0.03] | −0.03 [−0.31, 0.26] | 0.14 [−0.26, 0.53] |
| $\lambda_{10}$ | −0.30 [−0.58, −0.03] | 0.11 [−0.14, 0.36] | 0.05 [−0.38, 0.47] |
| $\lambda_{11}$ | 0.21 [−0.19, 0.62] | −0.06 [−0.25, 0.13] | 0.07 [−0.42, 0.55] |
| $\lambda_{12}$ | 0.28 [0.04, 0.51] | −0.17 [−0.41, 0.07] | 0.13 [−0.50, 0.76] |
| $\lambda_{13}$ | 0.03 [−0.14, 0.20] | 0.26 [−0.01, 0.53] | 0.13 [−0.04, 0.31] |
| $\lambda_{14}$ | 0.29 [0.08, 0.51] | 0.05 [−0.32, 0.43] | −0.04 [−0.42, 0.34] |

*Note.* We report standardized parameter estimates and 95.00 % CI intervals for all variables, except for the means of fixed/random effects; here, we report unstandardized estimates, as indicated by *.
Model fit: $\chi^2(972)$ = 2100.69 $p$ <.001; robust TLI = 0.944; robust CFI = 0.952; SRMR = 0.100; robust RMSEA [90% CI] = 0.076 [0.061, 0.089].

Supplementary Table 9: Multiple group analysis: Living alone (yes vs. no)

| | Yes $n$ = 281 | No $n$ = 145 |
|---|---|---|
| Parameter | Est.[95%CI] | Est.[95%CI] |
| Random effects: Means | | |
| Intercept FI1* | 0.19 [0.18, 0.21] | 0.16 [0.14, 0.18] |
| Intercept FI2* | 0.21 [0.20, 0.23] | 0.18 [0.16, 0.21] |
| Intercept LS1* | 3.48 [3.36, 3.60] | 3.24 [3.11, 3.37] |
| Intercept LS2* | 3.56 [3.43, 3.68] | 3.41 [3.27, 3.56] |
| Fixed effects: Means | | |
| Slope LS1* | 0.01 [−0.01, 0.03] | 0.02 [0.00, 0.04] |
| Slope LS2* | −0.01 [−0.02, 0.01] | −0.02 [−0.03, −0.00] |
| Random Effects: Correlation | | |
| $\zeta_1$: Intercept FI1 ↔ Intercept FI2 | 0.95 [0.92, 0.97] | 0.94 [0.89, 0.99] |
| $\zeta_2$: Intercept FI1 ↔ Intercept LS1 | 0.55 [0.42, 0.68] | 0.59 [0.39, 0.78] |
| $\zeta_3$: Intercept FI1 ↔ Intercept LS2 | 0.55 [0.41, 0.69] | 0.65 [0.46, 0.84] |
| $\zeta_4$: Intercept FI2 ↔ Intercept LS1 | 0.52 [0.38, 0.66] | 0.48 [0.24, 0.72] |
| $\zeta_5$: Intercept FI2 ↔ Intercept LS2 | 0.51 [0.36, 0.66] | 0.52 [0.28, 0.76] |
| $\zeta_6$: Intercept LS1 ↔ Intercept LS2 | 0.93 [0.86, 1.00] | 0.95 [0.84, 1.06] |
| Autoregressive (FI → FI) | | |
| $\alpha_1$ | 0.26 [0.07, 0.45] | 0.22 [−0.04, 0.48] |
| $\alpha_2$ | 0.26 [0.09, 0.43] | 0.37 [0.12, 0.63] |
| $\alpha_3$ | 0.29 [0.02, 0.55] | 0.28 [−0.21, 0.76] |
| $\alpha_4$ | 0.16 [−0.02, 0.33] | −0.22 [−0.79, 0.34] |
| $\alpha_5$ | 0.34 [0.15, 0.53] | 0.09 [−0.22, 0.40] |
| $\alpha_6$ | 0.24 [0.04, 0.44] | 0.18 [−0.15, 0.52] |
| $\alpha_7$ | −0.05 [−0.23, 0.13] | 0.36 [−0.03, 0.75] |
| $\alpha_8$ | 0.17 [−0.03, 0.37] | 0.27 [0.03, 0.50] |
| $\alpha_9$ | 0.09 [−0.19, 0.38] | 0.31 [−0.04, 0.66] |
| $\alpha_{10}$ | 0.11 [−0.31, 0.53] | 0.14 [−1.15, 1.43] |
| $\alpha_{11}$ | 0.23 [−0.02, 0.49] | 0.60 [0.30, 0.89] |
| $\alpha_{12}$ | 0.22 [−0.03, 0.46] | 0.44 [0.02, 0.85] |
| $\alpha_{13}$ | 0.15 [−0.12, 0.43] | 0.73 [0.51, 0.94] |
| Autoregressive (LS → LS) | | |
| $\beta_1$ | 0.25 [0.08, 0.41] | 0.22 [−0.16, 0.60] |
| $\beta_2$ | 0.24 [0.01, 0.48] | −0.08 [−0.57, 0.41] |
| $\beta_3$ | 0.08 [−0.23, 0.40] | 0.36 [−0.07, 0.79] |
| $\beta_4$ | 0.07 [−0.23, 0.37] | −0.12 [−0.73, 0.49] |
| $\beta_5$ | 0.09 [−0.19, 0.36] | 0.37 [−0.05, 0.78] |
| $\beta_6$ | 0.32 [0.13, 0.52] | 0.09 [−0.33, 0.52] |
| $\beta_7$ | 0.01 [−0.30, 0.32] | −0.04 [−0.65, 0.58] |
| $\beta_8$ | 0.11 [−0.16, 0.38] | 0.07 [−0.46, 0.61] |
| $\beta_9$ | 0.20 [−0.13, 0.53] | −0.28 [−0.87, 0.30] |
| $\beta_{10}$ | 0.01 [−0.24, 0.26] | 0.08 [−0.57, 0.72] |
| $\beta_{11}$ | −0.18 [−0.56, 0.20] | −0.10 [−0.48, 0.27] |
| $\beta_{12}$ | 0.02 [−0.32, 0.36] | −0.09 [−0.87, 0.69] |
| $\beta_{13}$ | 0.31 [0.00, 0.63] | 0.33 [−0.54, 1.20] |
| Cross-lagged (LS → FI) | | |
| $\delta_1$ | −0.06 [−0.21, 0.09] | 0.22 [−0.24, 0.68] |
| $\delta_2$ | 0.04 [−0.11, 0.18] | 0.27 [0.01, 0.53] |
| $\delta_3$ | 0.12 [−0.05, 0.29] | −0.24 [−0.53, 0.06] |
| $\delta_4$ | 0.01 [−0.18, 0.19] | −0.11 [−0.53, 0.31] |
| $\delta_5$ | −0.06 [−0.24, 0.11] | −0.05 [−0.44, 0.35] |
| $\delta_6$ | −0.02 [−0.18, 0.14] | −0.08 [−0.27, 0.12] |
| $\delta_7$ | 0.08 [−0.05, 0.21] | 0.20 [−0.14, 0.54] |
| $\delta_8$ | −0.12 [−0.29, 0.05] | 0.31 [−0.05, 0.67] |
| $\delta_9$ | −0.04 [−0.25, 0.17] | 0.37 [−0.15, 0.88] |
| $\delta_{10}$ | 0.01 [−0.22, 0.23] | −0.12 [−0.49, 0.26] |
| $\delta_{11}$ | 0.08 [−0.10, 0.26] | 0.15 [−0.15, 0.46] |
| $\delta_{12}$ | 0.05 [−0.20, 0.31] | 0.06 [−0.15, 0.28] |
| $\delta_{13}$ | 0.15 [−0.03, 0.34] | −0.23 [−0.50, 0.04] |
| Cross-lagged (FI → LS) | | |
| $\gamma_1$ | 0.12 [−0.01, 0.25] | 0.06 [−0.15, 0.28] |
| $\gamma_2$ | 0.13 [−0.05, 0.31] | 0.32 [−0.06, 0.69] |
| $\gamma_3$ | 0.16 [−0.04, 0.36] | 0.28 [−0.07, 0.62] |
| $\gamma_4$ | −0.09 [−0.32, 0.13] | 0.11 [−0.52, 0.74] |
| $\gamma_5$ | −0.03 [−0.25, 0.20] | −0.04 [−0.25, 0.17] |
| $\gamma_6$ | −0.08 [−0.22, 0.06] | −0.11 [−0.33, 0.12] |
| $\gamma_7$ | 0.05 [−0.10, 0.20] | 0.01 [−0.21, 0.24] |
| $\gamma_8$ | 0.05 [−0.16, 0.26] | 0.17 [−0.27, 0.62] |
| $\gamma_9$ | 0.01 [−0.22, 0.24] | −0.05 [−0.47, 0.37] |
| $\gamma_{10}$ | 0.06 [−0.19, 0.30] | 0.57 [−0.06, 1.20] |
| $\gamma_{11}$ | 0.28 [0.01, 0.55] | 0.23 [−0.43, 0.89] |
| $\gamma_{12}$ | 0.13 [−0.12, 0.39] | −0.29 [−1.11, 0.52] |
| $\gamma_{13}$ | 0.05 [−0.15, 0.25] | 0.35 [−0.21, 0.92] |
| Within-time (FI ↔ LS) | | |
| $\lambda_1$ | 0.22 [0.09, 0.35] | 0.29 [0.10, 0.48] |
| $\lambda_2$ | 0.17 [0.03, 0.32] | 0.16 [−0.03, 0.36] |
| $\lambda_3$ | 0.20 [0.08, 0.32] | 0.28 [0.02, 0.54] |
| $\lambda_4$ | 0.18 [−0.01, 0.37] | 0.10 [−0.19, 0.38] |
| $\lambda_5$ | 0.23 [−0.04, 0.50] | 0.05 [−0.42, 0.51] |
| $\lambda_6$ | 0.23 [0.04, 0.42] | −0.01 [−0.25, 0.22] |
| $\lambda_7$ | 0.25 [0.10, 0.40] | 0.05 [−0.18, 0.28] |
| $\lambda_8$ | 0.03 [−0.15, 0.21] | 0.28 [−0.02, 0.58] |
| $\lambda_9$ | 0.06 [−0.14, 0.27] | −0.01 [−0.48, 0.46] |
| $\lambda_{10}$ | 0.08 [−0.10, 0.27] | 0.15 [−0.44, 0.74] |
| $\lambda_{11}$ | 0.14 [−0.07, 0.36] | 0.38 [−0.22, 0.98] |
| $\lambda_{12}$ | 0.23 [−0.10, 0.57] | −0.21 [−0.45, 0.04] |
| $\lambda_{13}$ | 0.18 [0.02, 0.34] | −0.01 [−0.19, 0.18] |
| $\lambda_{14}$ | 0.08 [−0.09, 0.25] | 0.26 [−0.38, 0.89] |

*Note.* We report standardized parameter estimates and 95.00 % CI intervals for all variables, except for the means of fixed/random effects; here, we report unstandardized estimates, as indicated by *.

Model fit: $\chi^2(648) = 1633.09$ $p < .001$; robust TLI = 0.953; robust CFI = 0.960; SRMR = 0.082; robust RMSEA [90% CI] = 0.071 [0.057, 0.085].

Supplementary Table 10: Multiple group analysis: social participation (yes vs. no; participation in at least one social activity)

| | Yes $n$ = 249 | No $n$ = 177 |
|---|---|---|
| Parameter | Est.[95%CI] | Est.[95%CI] |
| Random effects: Means | | |
| Intercept FI1* | 0.14 [0.13, 0.16] | 0.21 [0.19, 0.23] |
| Intercept FI2* | 0.17 [0.15, 0.18] | 0.23 [0.21, 0.25] |
| Intercept LS1* | 3.28 [3.19, 3.38] | 3.47 [3.34, 3.61] |
| Intercept LS2* | 3.33 [3.22, 3.44] | 3.63 [3.49, 3.78] |
| Fixed effects: Means | | |
| Slope LS1* | 0.01 [−0.01, 0.02] | 0.02 [0.00, 0.04] |
| Slope LS2* | −0.00 [−0.02, 0.01] | −0.01 [−0.03, 0.01] |
| Random Effects: Correlation | | |
| $\zeta_1$: Intercept FI1 ↔ Intercept FI2 | 0.94 [0.90, 0.97] | 0.94 [0.90, 0.97] |
| $\zeta_2$: Intercept FI1 ↔ Intercept LS1 | 0.44 [0.27, 0.61] | 0.60 [0.48, 0.72] |
| $\zeta_3$: Intercept FI1 ↔ Intercept LS2 | 0.42 [0.25, 0.59] | 0.61 [0.48, 0.73] |
| $\zeta_4$: Intercept FI2 ↔ Intercept LS1 | 0.37 [0.16, 0.58] | 0.55 [0.42, 0.67] |
| $\zeta_5$: Intercept FI2 ↔ Intercept LS2 | 0.37 [0.14, 0.59] | 0.57 [0.44, 0.70] |
| $\zeta_6$: Intercept LS1 ↔ Intercept LS2 | 0.95 [0.87, 1.03] | 0.91 [0.84, 0.98] |
| Autoregressive (FI → FI) | | |
| $\alpha_1$ | 0.31 [0.08, 0.53] | 0.23 [0.02, 0.43] |
| $\alpha_2$ | 0.15 [−0.02, 0.32] | 0.36 [0.17, 0.55] |
| $\alpha_3$ | 0.17 [−0.04, 0.37] | 0.34 [0.01, 0.68] |
| $\alpha_4$ | −0.06 [−0.37, 0.26] | 0.15 [−0.07, 0.37] |
| $\alpha_5$ | 0.33 [0.09, 0.56] | 0.30 [0.06, 0.55] |
| $\alpha_6$ | 0.25 [−0.01, 0.50] | 0.24 [0.02, 0.46] |
| $\alpha_7$ | 0.31 [−0.18, 0.80] | 0.01 [−0.25, 0.26] |
| $\alpha_8$ | 0.22 [−0.06, 0.50] | 0.20 [0.02, 0.39] |
| $\alpha_9$ | 0.27 [−0.04, 0.57] | 0.08 [−0.14, 0.31] |
| $\alpha_{10}$ | 0.29 [−0.22, 0.80] | −0.11 [−0.50, 0.28] |
| $\alpha_{11}$ | 0.31 [−0.05, 0.67] | 0.29 [0.01, 0.57] |
| $\alpha_{12}$ | 0.24 [−0.07, 0.55] | 0.23 [−0.01, 0.47] |
| $\alpha_{13}$ | 0.36 [0.03, 0.69] | 0.35 [0.04, 0.67] |
| Autoregressive (LS → LS) | | |
| $\beta_1$ | 0.15 [−0.14, 0.44] | 0.26 [0.09, 0.44] |
| $\beta_2$ | 0.30 [0.05, 0.55] | 0.10 [−0.19, 0.38] |
| $\beta_3$ | 0.29 [0.01, 0.58] | 0.05 [−0.32, 0.42] |
| $\beta_4$ | 0.04 [−0.19, 0.27] | 0.02 [−0.30, 0.35] |
| $\beta_5$ | 0.12 [−0.07, 0.30] | 0.14 [−0.17, 0.45] |
| $\beta_6$ | 0.38 [0.16, 0.60] | 0.22 [−0.03, 0.46] |
| $\beta_7$ | 0.36 [0.04, 0.68] | −0.24 [−0.52, 0.04] |
| $\beta_8$ | −0.07 [−0.47, 0.34] | 0.08 [−0.21, 0.36] |
| $\beta_9$ | 0.12 [−0.37, 0.62] | 0.02 [−0.38, 0.42] |
| $\beta_{10}$ | 0.27 [−0.20, 0.73] | −0.11 [−0.38, 0.16] |
| $\beta_{11}$ | −0.01 [−0.43, 0.41] | −0.16 [−0.51, 0.20] |
| $\beta_{12}$ | 0.27 [−0.18, 0.72] | −0.11 [−0.45, 0.24] |
| $\beta_{13}$ | 0.23 [−0.08, 0.53] | 0.52 [0.24, 0.80] |
| Cross-lagged (LS → FI) | | |
| $\delta_1$ | −0.13 [−0.32, 0.07] | 0.06 [−0.16, 0.28] |
| $\delta_2$ | 0.15 [−0.02, 0.32] | 0.06 [−0.11, 0.23] |
| $\delta_3$ | 0.02 [−0.20, 0.25] | 0.05 [−0.16, 0.25] |
| $\delta_4$ | −0.06 [−0.25, 0.13] | −0.04 [−0.24, 0.16] |
| $\delta_5$ | −0.01 [−0.21, 0.19] | −0.08 [−0.26, 0.10] |
| $\delta_6$ | −0.20 [−0.36, −0.03] | 0.03 [−0.13, 0.19] |
| $\delta_7$ | 0.13 [−0.06, 0.32] | 0.02 [−0.16, 0.21] |
| $\delta_8$ | −0.07 [−0.25, 0.11] | 0.05 [−0.17, 0.27] |
| $\delta_9$ | −0.03 [−0.27, 0.21] | 0.07 [−0.27, 0.40] |
| $\delta_{10}$ | 0.02 [−0.18, 0.22] | −0.05 [−0.32, 0.23] |
| $\delta_{11}$ | 0.12 [−0.14, 0.38] | 0.09 [−0.13, 0.31] |
| $\delta_{12}$ | 0.19 [−0.08, 0.45] | −0.04 [−0.22, 0.14] |
| $\delta_{13}$ | 0.16 [−0.07, 0.39] | 0.02 [−0.21, 0.24] |
| Cross-lagged (FI → LS) | | |
| $\gamma_1$ | 0.10 [−0.05, 0.25] | 0.12 [−0.01, 0.25] |
| $\gamma_2$ | 0.03 [−0.14, 0.19] | 0.21 [−0.04, 0.46] |
| $\gamma_3$ | 0.23 [0.06, 0.41] | 0.12 [−0.14, 0.38] |
| $\gamma_4$ | −0.15 [−0.34, 0.03] | −0.06 [−0.31, 0.19] |
| $\gamma_5$ | −0.11 [−0.30, 0.08] | −0.03 [−0.29, 0.22] |
| $\gamma_6$ | −0.05 [−0.21, 0.11] | −0.10 [−0.27, 0.07] |
| $\gamma_7$ | 0.01 [−0.21, 0.22] | 0.08 [−0.08, 0.24] |
| $\gamma_8$ | 0.13 [−0.28, 0.54] | 0.12 [−0.10, 0.33] |
| $\gamma_9$ | 0.10 [−0.14, 0.34] | −0.08 [−0.33, 0.16] |
| $\gamma_{10}$ | 0.02 [−0.19, 0.22] | 0.18 [−0.25, 0.60] |
| $\gamma_{11}$ | 0.19 [−0.19, 0.57] | 0.22 [−0.02, 0.46] |
| $\gamma_{12}$ | −0.11 [−0.32, 0.10] | 0.25 [−0.07, 0.58] |
| $\gamma_{13}$ | 0.30 [−0.04, 0.64] | 0.03 [−0.12, 0.18] |
| Within-time (FI ↔ LS) | | |
| $\lambda_1$ | 0.19 [−0.01, 0.38] | 0.26 [0.12, 0.40] |
| $\lambda_2$ | 0.24 [0.05, 0.43] | 0.14 [−0.01, 0.28] |
| $\lambda_3$ | 0.11 [−0.01, 0.23] | 0.20 [0.04, 0.36] |
| $\lambda_4$ | 0.19 [0.01, 0.37] | 0.13 [−0.11, 0.37] |
| $\lambda_5$ | 0.08 [−0.21, 0.37] | 0.17 [−0.10, 0.43] |
| $\lambda_6$ | 0.19 [0.02, 0.37] | 0.16 [−0.08, 0.41] |
| $\lambda_7$ | 0.27 [0.08, 0.47] | 0.18 [0.03, 0.33] |
| $\lambda_8$ | −0.02 [−0.31, 0.26] | 0.11 [−0.07, 0.29] |
| $\lambda_9$ | 0.06 [−0.18, 0.30] | 0.03 [−0.22, 0.29] |
| $\lambda_{10}$ | −0.11 [−0.28, 0.06] | 0.15 [−0.12, 0.42] |
| $\lambda_{11}$ | 0.09 [−0.16, 0.34] | 0.15 [−0.16, 0.47] |
| $\lambda_{12}$ | −0.02 [−0.28, 0.24] | 0.17 [−0.21, 0.55] |
| $\lambda_{13}$ | 0.04 [−0.13, 0.21] | 0.13 [−0.04, 0.31] |
| $\lambda_{14}$ | 0.12 [−0.16, 0.40] | 0.08 [−0.09, 0.25] |

*Note*. We report standardized parameter estimates and 95.00 % CI intervals for all variables, except for the means of fixed/random effects; here, we report unstandardized estimates, as indicated by *.

Model fit: $\chi^2(648)$ = 1343.61 $p$ <.001; robust TLI = 0.953; robust CFI = 0.960; SRMR = 0.065; robust RMSEA [90% CI] = 0.069 [0.058, 0.058].

Supplementary Table 11: Analysis with `bedrest` as time-varying covariate ($N = 162$)

| Parameter | Est.[95%CI] |
|---|---|
| Fixed/Random effects: Means | |
| Intercept FI1* | 0.16 [0.14, 0.17] |
| Intercept FI2* | 0.18 [0.16, 0.19] |
| Intercept LS1* | 3.32 [3.19, 3.45] |
| Intercept LS2* | 3.53 [3.37, 3.68] |
| Random effects: Means | |
| Slope LS1* | 0.02 [−0.01, 0.05] |
| Slope LS2* | −0.02 [−0.03, −0.01] |
| Random Effects: Correlation | |
| Intercept FI1 ↔ Intercept FI2 | 0.96 [0.93, 0.98] |
| Intercept FI1 ↔ Intercept LS1 | 0.50 [0.30, 0.70] |
| Intercept FI1 ↔ Intercept LS2 | 0.52 [0.35, 0.69] |
| Intercept FI2 ↔ Intercept LS1 | 0.51 [0.31, 0.70] |
| Intercept FI2 ↔ Intercept LS2 | 0.53 [0.38, 0.69] |
| Intercept LS1 ↔ Intercept LS2 | 0.97 [0.88, 1.07] |
| Autoregressive (FI → FI) | |
| $\alpha_1$ | 0.18 [0.01, 0.35] |
| $\alpha_2$ | 0.21 [−0.01, 0.43] |
| $\alpha_3$ | 0.27 [−0.02, 0.57] |
| $\alpha_4$ | 0.14 [−0.16, 0.44] |
| $\alpha_5$ | 0.27 [0.05, 0.49] |
| $\alpha_6$ | 0.21 [−0.06, 0.48] |
| $\alpha_7$ | −0.02 [−0.23, 0.20] |
| $\alpha_8$ | 0.15 [−0.09, 0.39] |
| $\alpha_9$ | 0.34 [0.17, 0.52] |
| $\alpha_{10}$ | 0.09 [−0.14, 0.33] |
| $\alpha_{11}$ | 0.09 [−0.09, 0.27] |
| $\alpha_{12}$ | 0.17 [−0.04, 0.39] |
| $\alpha_{13}$ | 0.40 [0.16, 0.65] |
| Autoregressive (LS → LS) | |
| $\beta_1$ | 0.28 [0.01, 0.55] |
| $\beta_2$ | 0.51 [0.27, 0.76] |
| $\beta_3$ | 0.46 [0.14, 0.78] |
| $\beta_4$ | 0.47 [0.16, 0.79] |
| $\beta_5$ | 0.41 [0.16, 0.66] |
| $\beta_6$ | 0.36 [0.13, 0.59] |
| $\beta_7$ | −0.07 [−0.42, 0.28] |
| $\beta_8$ | −0.15 [−0.44, 0.14] |
| $\beta_9$ | 0.06 [−0.36, 0.48] |
| $\beta_{10}$ | −0.20 [−0.51, 0.12] |
| $\beta_{11}$ | −0.23 [−0.49, 0.03] |
| $\beta_{12}$ | −0.04 [−0.34, 0.25] |
| $\beta_{13}$ | 0.22 [−0.08, 0.52] |
| Cross-lagged (LS → FI) | |
| $\delta_1$ | −0.05 [−0.22, 0.12] |
| $\delta_2$ | 0.12 [−0.03, 0.27] |
| $\delta_3$ | 0.07 [−0.13, 0.26] |
| $\delta_4$ | 0.04 [−0.22, 0.30] |
| $\delta_5$ | 0.03 [−0.16, 0.22] |
| $\delta_6$ | −0.00 [−0.25, 0.24] |
| $\delta_7$ | 0.08 [−0.10, 0.27] |
| $\delta_8$ | 0.09 [−0.25, 0.43] |
| $\delta_9$ | −0.01 [−0.22, 0.20] |
| $\delta_{10}$ | 0.06 [−0.11, 0.22] |
| $\delta_{11}$ | 0.06 [−0.10, 0.22] |
| $\delta_{12}$ | −0.04 [−0.20, 0.11] |
| $\delta_{13}$ | 0.09 [−0.05, 0.24] |
| Cross-lagged (FI → LS) | |
| $\gamma_1$ | 0.04 [−0.11, 0.19] |
| $\gamma_2$ | 0.20 [−0.03, 0.43] |
| $\gamma_3$ | 0.24 [0.02, 0.45] |
| $\gamma_4$ | −0.21 [−0.40, −0.01] |
| $\gamma_5$ | 0.15 [−0.05, 0.36] |
| $\gamma_6$ | 0.02 [−0.12, 0.16] |
| $\gamma_7$ | 0.15 [−0.11, 0.42] |
| $\gamma_8$ | −0.27 [−0.49, −0.05] |
| $\gamma_9$ | 0.08 [−0.19, 0.35] |
| $\gamma_{10}$ | 0.11 [−0.17, 0.39] |
| $\gamma_{11}$ | 0.13 [−0.07, 0.32] |
| $\gamma_{12}$ | −0.07 [−0.32, 0.19] |
| $\gamma_{13}$ | 0.12 [−0.07, 0.30] |
| Within-time (FI ↔ LS) | |
| $\lambda_1$ | 0.16 [−0.02, 0.34] |
| $\lambda_2$ | 0.10 [−0.09, 0.29] |
| $\lambda_3$ | 0.10 [−0.02, 0.23] |
| $\lambda_4$ | 0.25 [−0.08, 0.57] |
| $\lambda_5$ | −0.11 [−0.34, 0.11] |
| $\lambda_6$ | 0.27 [0.09, 0.44] |
| $\lambda_7$ | 0.16 [−0.05, 0.38] |
| $\lambda_8$ | −0.08 [−0.30, 0.14] |
| $\lambda_9$ | 0.17 [−0.09, 0.43] |
| $\lambda_{10}$ | 0.10 [−0.10, 0.31] |
| $\lambda_{11}$ | −0.11 [−0.35, 0.12] |
| $\lambda_{12}$ | 0.15 [−0.19, 0.49] |
| $\lambda_{13}$ | 0.03 [−0.14, 0.20] |
| $\lambda_{14}$ | 0.09 [−0.06, 0.23] |
| Time-varying covariate | |
| $\text{bedrest}_1 \rightarrow \text{r\_fi}_1$ | 0.21 [0.06, 0.35] |
| $\text{bedrest}_1 \rightarrow \text{r\_ucla}_1$ | −0.08 [−0.21, 0.04] |
| $\text{bedrest}_2 \rightarrow \text{r\_fi}_2$ | 0.42 [0.23, 0.60] |
| $\text{bedrest}_2 \rightarrow \text{r\_ucla}_2$ | 0.00 [−0.20, 0.20] |
| $\text{bedrest}_3 \rightarrow \text{r\_fi}_3$ | 0.32 [0.03, 0.60] |
| $\text{bedrest}_3 \rightarrow \text{r\_ucla}_3$ | −0.13 [−0.29, 0.02] |
| $\text{bedrest}_4 \rightarrow \text{r\_fi}_4$ | 0.28 [0.10, 0.46] |
| $\text{bedrest}_4 \rightarrow \text{r\_ucla}_4$ | −0.09 [−0.27, 0.09] |
| $\text{bedrest}_5 \rightarrow \text{r\_fi}_5$ | 0.39 [0.20, 0.58] |
| $\text{bedrest}_5 \rightarrow \text{r\_ucla}_5$ | 0.18 [−0.00, 0.37] |
| $\text{bedrest}_6 \rightarrow \text{r\_fi}_6$ | 0.29 [0.13, 0.46] |
| $\text{bedrest}_6 \rightarrow \text{r\_ucla}_6$ | 0.04 [−0.08, 0.17] |
| $\text{bedrest}_7 \rightarrow \text{r\_fi}_7$ | 0.24 [0.03, 0.46] |
| $\text{bedrest}_7 \rightarrow \text{r\_ucla}_7$ | −0.02 [−0.14, 0.11] |
| $\text{bedrest}_8 \rightarrow \text{r\_fi}_8$ | 0.37 [0.18, 0.56] |
| $\text{bedrest}_8 \rightarrow \text{r\_ucla}_8$ | −0.12 [−0.28, 0.05] |
| $\text{bedrest}_9 \rightarrow \text{r\_fi}_9$ | 0.30 [0.11, 0.50] |
| $\text{bedrest}_9 \rightarrow \text{r\_ucla}_9$ | 0.23 [0.05, 0.42] |
| $\text{bedrest}_{10} \rightarrow \text{r\_fi}_{10}$ | 0.33 [0.20, 0.47] |
| $\text{bedrest}_{10} \rightarrow \text{r\_ucla}_{10}$ | −0.13 [−0.26, 0.00] |
| $\text{bedrest}_{11} \rightarrow \text{r\_fi}_{11}$ | 0.20 [0.02, 0.39] |
| $\text{bedrest}_{11} \rightarrow \text{r\_ucla}_{11}$ | 0.09 [−0.08, 0.27] |
| $\text{bedrest}_{12} \rightarrow \text{r\_fi}_{12}$ | 0.34 [0.15, 0.52] |
| $\text{bedrest}_{12} \rightarrow \text{r\_ucla}_{12}$ | 0.01 [−0.22, 0.23] |
| $\text{bedrest}_{13} \rightarrow \text{r\_fi}_{13}$ | 0.26 [0.11, 0.42] |
| $\text{bedrest}_{13} \rightarrow \text{r\_ucla}_{13}$ | 0.19 [−0.01, 0.39] |
| $\text{bedrest}_{14} \rightarrow \text{r\_fi}_{14}$ | 0.28 [0.12, 0.45] |
| $\text{bedrest}_{14} \rightarrow \text{r\_ucla}_{14}$ | 0.06 [−0.15, 0.27] |

*Note.* We report standardized parameter estimates and 95.00 % CI intervals for all variables, except for the means of fixed/random effects; here, we report unstandardized estimates, as indicated by *. Model fit: $\chi^2(688) = 1195.14$ $p < .001$; robust TLI = 0.942; robust CFI = 0.948; SRMR = 0.148; robust RMSEA [90% CI] = 0.055 [0.047, 0.064].

Supplementary Table 12: Analysis with `falls` as time-varying covariate ($N = 162$)

| Parameters | Est. [95%CI] |
|---|---|
| Random effects: Means | |
| Intercept FI1* | 0.16 [0.15, 0.18] |
| Intercept FI2* | 0.18 [0.16, 0.20] |
| Intercept LS1* | 3.32 [3.19, 3.44] |
| Intercept LS2* | 3.53 [3.38, 3.68] |
| Fixed effects: Means | |
| Slope LS1* | 0.02 [−0.00, 0.05] |
| Slope LS2* | −0.02 [−0.03, −0.00] |
| Random Effects: Correlation | |
| Intercept FI1 ↔ Intercept FI2 | 0.96 [0.93, 0.99] |
| Intercept FI1 ↔ Intercept LS1 | 0.52 [0.30, 0.74] |
| Intercept FI1 ↔ Intercept LS2 | 0.54 [0.37, 0.71] |
| Intercept FI2 ↔ Intercept LS1 | 0.52 [0.33, 0.71] |
| Intercept FI2 ↔ Intercept LS2 | 0.55 [0.40, 0.71] |
| Intercept LS1 ↔ Intercept LS2 | 0.95 [0.84, 1.06] |
| Autoregressive (FI → FI) | |
| $\alpha_1$ | 0.24 [0.04, 0.43] |
| $\alpha_2$ | 0.30 [0.04, 0.56] |
| $\alpha_3$ | 0.37 [0.04, 0.70] |
| $\alpha_4$ | 0.18 [−0.20, 0.55] |
| $\alpha_5$ | 0.22 [−0.07, 0.51] |
| $\alpha_6$ | 0.19 [−0.08, 0.46] |
| $\alpha_7$ | -0.09 [−0.34, 0.16] |
| $\alpha_8$ | 0.12 [−0.14, 0.38] |
| $\alpha_9$ | 0.38 [0.17, 0.59] |
| $\alpha_{10}$ | 0.09 [−0.18, 0.36] |
| $\alpha_{11}$ | 0.18 [−0.02, 0.38] |
| $\alpha_{12}$ | 0.24 [0.02, 0.46] |
| $\alpha_{13}$ | 0.48 [0.23, 0.73] |
| Autoregressive (LS → LS) | |
| $\beta_1$ | 0.26 [−0.01, 0.54] |
| $\beta_2$ | 0.49 [0.23, 0.76] |
| $\beta_3$ | 0.44 [0.07, 0.80] |
| $\beta_4$ | 0.45 [0.12, 0.79] |
| $\beta_5$ | 0.40 [0.13, 0.66] |
| $\beta_6$ | 0.35 [0.11, 0.58] |
| $\beta_7$ | −0.09 [−0.44, 0.26] |
| $\beta_8$ | −0.19 [−0.49, 0.11] |
| $\beta_9$ | 0.05 [−0.39, 0.49] |
| $\beta_{10}$ | −0.21 [−0.53, 0.11] |
| $\beta_{11}$ | −0.24 [−0.53, 0.05] |
| $\beta_{12}$ | −0.01 [−0.30, 0.29] |
| $\beta_{13}$ | 0.24 [−0.07, 0.55] |
| Cross-lagged (LS → FI) | |
| $\delta_1$ | −0.09 [−0.27, 0.10] |
| $\delta_2$ | 0.12 [−0.01, 0.24] |
| $\delta_3$ | 0.05 [−0.15, 0.25] |
| $\delta_4$ | 0.07 [−0.21, 0.34] |
| $\delta_5$ | 0.02 [−0.21, 0.26] |
| $\delta_6$ | −0.06 [−0.34, 0.22] |
| $\delta_7$ | 0.07 [−0.12, 0.26] |
| $\delta_8$ | 0.07 [−0.27, 0.41] |
| $\delta_9$ | −0.01 [−0.23, 0.22] |
| $\delta_{10}$ | 0.03 [−0.17, 0.23] |
| $\delta_{11}$ | 0.14 [−0.06, 0.34] |
| $\delta_{12}$ | −0.01 [−0.17, 0.15] |
| $\delta_{13}$ | 0.13 [−0.00, 0.25] |
| Cross-lagged (FI → LS) | |
| $\gamma_1$ | 0.05 [−0.12, 0.22] |
| $\gamma_2$ | 0.19 [−0.02, 0.40] |
| $\gamma_3$ | 0.21 [−0.05, 0.46] |
| $\gamma_4$ | −0.23 [−0.46, −0.00] |
| $\gamma_5$ | 0.07 [−0.18, 0.32] |
| $\gamma_6$ | 0.01 [−0.12, 0.14] |
| $\gamma_7$ | 0.20 [−0.07, 0.46] |
| $\gamma_8$ | −0.24 [−0.50, 0.02] |
| $\gamma_9$ | 0.04 [−0.23, 0.30] |
| $\gamma_{10}$ | 0.09 [−0.19, 0.37] |
| $\gamma_{11}$ | 0.10 [−0.09, 0.29] |
| $\gamma_{12}$ | −0.04 [−0.30, 0.23] |
| $\gamma_{13}$ | 0.14 [−0.03, 0.31] |
| Within-time (FI ↔ LS) | |
| $\lambda_1$ | 0.16 [−0.03, 0.35] |
| $\lambda_2$ | 0.09 [−0.11, 0.29] |
| $\lambda_3$ | 0.06 [−0.09, 0.20] |
| $\lambda_4$ | 0.20 [−0.15, 0.54] |
| $\lambda_5$ | −0.06 [−0.28, 0.17] |
| $\lambda_6$ | 0.24 [0.06, 0.42] |
| $\lambda_7$ | 0.14 [−0.09, 0.37] |
| $\lambda_8$ | −0.09 [−0.32, 0.14] |
| $\lambda_9$ | 0.22 [−0.03, 0.47] |
| $\lambda_{10}$ | 0.04 [−0.17, 0.25] |
| $\lambda_{11}$ | −0.14 [−0.39, 0.10] |
| $\lambda_{12}$ | 0.15 [−0.21, 0.52] |
| $\lambda_{13}$ | 0.11 [−0.05, 0.26] |
| $\lambda_{14}$ | 0.10 [−0.02, 0.22] |
| Time-varying covariate | |
| $\text{falls}_1 \rightarrow \text{r\_fi}_1$ | 0.34 [0.10, 0.59] |
| $\text{falls}_1 \rightarrow \text{r\_ucla}_1$ | 0.01 [−0.07, 0.09] |
| $\text{falls}_2 \rightarrow \text{r\_fi}_2$ | 0.16 [−0.09, 0.41] |
| $\text{falls}_2 \rightarrow \text{r\_ucla}_2$ | −0.02 [−0.06, 0.01] |
| $\text{falls}_3 \rightarrow \text{r\_fi}_3$ | −0.04 [−0.08, −0.00] |
| $\text{falls}_3 \rightarrow \text{r\_ucla}_3$ | −0.11 [−0.22, 0.00] |
| $\text{falls}_4 \rightarrow \text{r\_fi}_4$ | 0.09 [−0.12, 0.31] |
| $\text{falls}_4 \rightarrow \text{r\_ucla}_4$ | −0.11 [−0.30, 0.08] |
| $\text{falls}_5 \rightarrow \text{r\_fi}_5$ | −0.04 [−0.11, 0.03] |
| $\text{falls}_5 \rightarrow \text{r\_ucla}_5$ | −0.02 [−0.04, −0.00] |
| $\text{falls}_6 \rightarrow \text{r\_fi}_6$ | 0.08 [−0.01, 0.18] |
| $\text{falls}_6 \rightarrow \text{r\_ucla}_6$ | 0.00 [−0.14, 0.14] |
| $\text{falls}_7 \rightarrow \text{r\_fi}_7$ | 0.02 [−0.15, 0.19] |
| $\text{falls}_7 \rightarrow \text{r\_ucla}_7$ | −0.05 [−0.09, −0.00] |
| $\text{falls}_8 \rightarrow \text{r\_fi}_8$ | 0.13 [−0.06, 0.33] |
| $\text{falls}_8 \rightarrow \text{r\_ucla}_8$ | −0.02 [−0.14, 0.09] |
| $\text{falls}_9 \rightarrow \text{r\_fi}_9$ | 0.29 [−0.09, 0.67] |
| $\text{falls}_9 \rightarrow \text{r\_ucla}_9$ | −0.03 [−0.09, 0.02] |
| $\text{falls}_{10} \rightarrow \text{r\_fi}_{10}$ | 0.04 [−0.14, 0.23] |
| $\text{falls}_{10} \rightarrow \text{r\_ucla}_{10}$ | −0.09 [−0.17, −0.00] |
| $\text{falls}_{11} \rightarrow \text{r\_fi}_{11}$ | 0.07 [−0.03, 0.17] |
| $\text{falls}_{11} \rightarrow \text{r\_ucla}_{11}$ | −0.00 [−0.03, 0.02] |
| $\text{falls}_{12} \rightarrow \text{r\_fi}_{12}$ | 0.12 [−0.03, 0.28] |
| $\text{falls}_{12} \rightarrow \text{r\_ucla}_{12}$ | −0.02 [−0.07, 0.02] |
| $\text{falls}_{13} \rightarrow \text{r\_fi}_{13}$ | 0.15 [−0.01, 0.31] |
| $\text{falls}_{13} \rightarrow \text{r\_ucla}_{13}$ | −0.08 [−0.19, 0.03] |
| $\text{falls}_{14} \rightarrow \text{r\_fi}_{14}$ | 0.05 [0.03, 0.07] |
| $\text{falls}_{14} \rightarrow \text{r\_ucla}_{14}$ | 0.07 [0.03, 0.11] |

*Note.* We report standardized parameter estimates and 95.00 % CI intervals for all variables, except for the means of fixed/random effects; here, we report unstandardized estimates, as indicated by *. Model fit: $\chi^2(688) = 1035.99$ $p < .001$; robust TLI = 0.947; robust CFI = 0.953; SRMR = 0.070; robust RMSEA [90% CI] = 0.052 [0.044, 0.059].

#### Supplementary References


Curran, P. J., Howard, A. L., Bainter, S. A., Lane, S. T., & McGinley, J. S. (2014). The separation of between-person and within-person components of individual change over time: A latent curve model with structured residuals. *Journal of consulting and clinical psychology*, *82*(5), 879. https://doi.org/10.1037/a0035297

McNeish, D., & Matta, T. (2018). Differentiating between mixed-effects and latent-curve approaches to growth modeling. *Behavior research methods*, *50*, 1398–1414. https://doi.org/10.3758/s13428-017-0976-5

Stolz, E. (2024). *FRequent health Assessment In Later life (FRAIL70+) (SUF edition)*. https://doi.org/10.11587/DJNOHX
